# Personalised approach to hypertension treatment: Rationale and design of the HYPERMARKER randomised trial

**DOI:** 10.64898/2026.02.04.26345342

**Authors:** Matthew Chapman, Jonathan E. Knikman, Alastair Mobley, Bart Lagerwaard, Fernando Martinez-Garcia, Seraphine Zeitouny, Daniel Engler, Renate B. Schnabel, Wilko Spiering, Iris D. Bos, A Champsi, Alexander W. Carter, Thomas Hankemeier, Diederick Grobbee, Dipak Kotecha, the HYPERMARKER Consortium

## Abstract

**Background and Objective:** Blood pressure treatment response is variable in individual patients, and the choice of medical therapy is often dependent on clinician experience. Treatment choices can be personalised by patient empowerment, metabolomic profiles and augmented by machine learning, but robust evaluation is lacking on how these can be combined to enhance clinical effectiveness. The HYPERMARKER trial will evaluate how an individualised choice of medication class can address the avoidable global health and economic burdens of hypertension.

**Design and Intervention:** The HYPERMARKER trial is a proof-of-concept, pragmatic, multicentre, adaptive, open-label strategy trial embedded into routine clinical practice with stratified individual patient randomisation. The trial was co-designed with a patient and public involvement team. The intervention is a digital portal that supports shared decision making on hypertension therapy class using clinical features plus metabolomic profiles determined with liquid chromatography-mass spectrometry.

**Participants and Outcomes:** Eligible patients are aged ≥18yrs with a systolic blood pressure ≥140mmHg and a clinical indication for antihypertensive therapy. 400 participants will be randomised to usual standard of care for treatment selection, or to the intervention group. Remote follow-up will occur through a patient smartphone application and linked blood pressure monitor to assess the primary outcome of change in home systolic blood pressure during a four-week period after medication changes. Secondary outcomes will include patient-reported adverse effects and quality of life, treatment withdrawal, healthcare utilisation and a health economic analysis. In the second phase of the trial, all participants will receive an updated version of the intervention, regardless of original randomised group.

**Ethics and Dissemination:** Ethical approval will be obtained for all sites. Approval in England: North West – Greater Manchester West Research Ethics Committee (REC) (25/NW/0296). Trial results will be disseminated via peer-reviewed publications and plain language patient summaries.

**Trial registration:** Clinicaltrials.gov: NCT07294794; ISRCTN: ISRCTN29385951.

**STRENGTH AND LIMITATIONS OF THIS STUDY:**

- Hypertension is a major cause of preventable morbidity and mortality, and this study aims to reduce those burdens through machine learning-based integration of metabolomics with clinical factors to enable better personalisation of blood pressure lowering therapy.
- The HYPERMARKER trial was co-created with patient and public representatives, using digital technology with remote monitoring to facilitate a high level of shared care and patient empowerment.
- HYPERMARKER is a pragmatic proof-of-concept randomised trial designed to evaluate the potential for future pharmacometabolomic strategies to aid clinical decision-making.

## INTRODUCTION

Hypertension affects over 30% of adults worldwide, with global prevalence having doubled between 1990 and 2019.^[1,2]^ Hypertension is a leading modifiable risk factor for cardiovascular, cerebrovascular and renal disease, leading to substantial morbidity and mortality.^[3]^ Despite being readily identifiable and treatable, it is often poorly managed. Globally, only 23% of women and 18% of men with hypertension achieved blood pressure levels within recommended target ranges in 2019.^[1]^ As a consequence, hypertension has a major economic impact, which could be substantially reduced through improving the use of existing drug therapy.^[4]^ Regardless of other patient comorbidities, reducing blood pressure has a significant patient benefit, including around 10% lower risk of major cardiovascular events for every 5mmHg drop in systolic blood pressure (SBP).^[5]^

While the management of hypertension may be multifaceted and include lifestyle interventions, most patients require long-term drug therapy. Current European and American guidelines advocate the use of initial low dose dual combination therapy, using either a thiazide diuretic, calcium channel blocker, angiotensin converting enzyme inhibitor (ACEi) or angiotensin receptor blocker (ARB).^[6,7]^ However, limited data exist to guide clinicians in their choice for each individual patient. Where recommendations are made, this often relates to cardiovascular comorbidities where antihypertensives may have beneficial effect outside of blood pressure lowering.^[8]^ One year after initiation, adherence to blood pressure medication is less than 50%,^[9]^ highlighting how adherence and side effect profiles are critical to overall management^[10,11]^ and the need to consider each patient individually.^[12]^

### CURRENT CLINICAL PRACTICE IN HYPERTENSION MANAGEMENT

To gain real-word perspectives on hypertension management, we pooled data on the management of hypertension across five large studies using electronic healthcare records. In total, 6,334,007 patients with hypertension were included from the United Kingdom, United States, Germany, Italy and Canada, enrolled between 1996 and 2019.^[13–17]^ The drug classes used as initial therapy for study-defined hypertension were: ACEi/ARB 43.3%; thiazide diuretics 18.8%; calcium channel blockers 15%; beta-blockers 15.4%; and other therapies 7.5% ***(Figure 1A)***. Four of the five studies (n=4,693,178 patients) reported whether the initial therapy used a single agent or combination therapy, with combinations used in 32.1%. These figures represent a clear variation from current international guideline recommendations.

**Figure 1.**
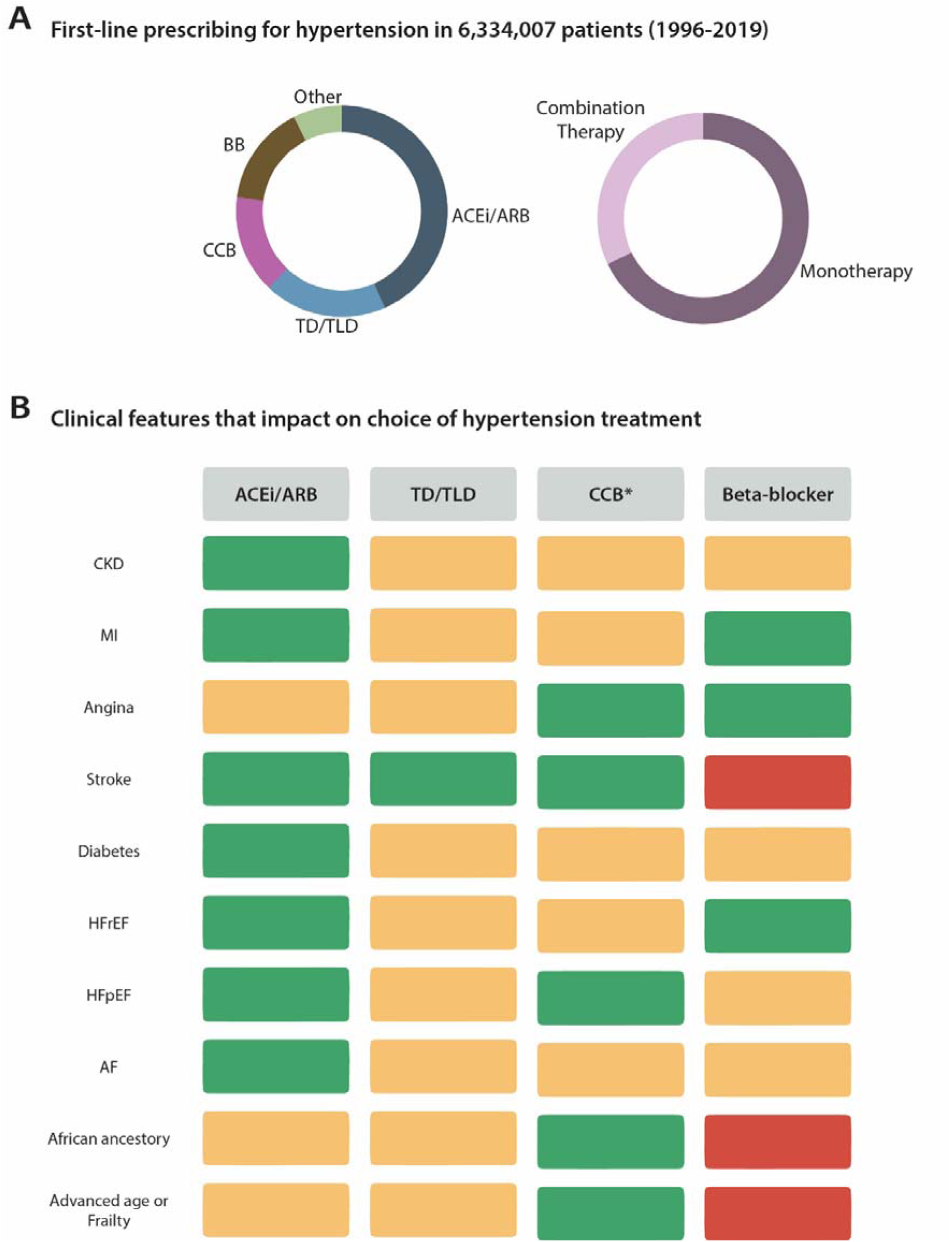
Choice of blood pressure lowering treatment class in routine practice. (A) Pooled data from 6,334,007 patients enrolled between 1996 and 2019 across five studies using electronic healthcare records from the United Kingdom, United States, Germany, Italy and Canada^[13–17]^. The drug classes used as initial therapy for study-defined hypertension were: ACEi/ARB 43.3%; thiazide diuretics 18.8%; calcium channel blockers 15%; beta-blockers 15.4%; and other therapies 7.5%. Four of the five studies reported whether initial therapy was a single agent (67.9%) or combination therapy (32.1%) (n=4,693,178 patients). (B) Clinical factors that may impact the most commonly prescribed drug classes for hypertension in clinical practice. Green, often used as first-line for hypertension management or another prognostic indication; amber, used in clinical practice but may not be first line; red, typically avoided in clinical practice. *refers to dihydropyridine calcium channel blockers. *BB, Beta-blockers; ACEi, Angiotensin converting enzyme inhibitors; ARB Angiotensin II receptor blockers; TD, Thiazide diuretic; TLD, Thiazide-like diuretic; CCB, Calcium channel blocker; CKD, Chronic Kidney Disease; MI, Myocardial infarction; HFrEF, Heart failure with reduced ejection fraction; AF, Atrial fibrillation*.

Reviewing consensus documents, guidelines and usual clinical practice, it is apparent that whilst there are several comorbid conditions that often dictate which drug class is used in a particular patient ***(Figure 1B)***, the ability to personalise blood pressure treatment remains significantly limited in clinical practice. Moreover, these comorbid conditions are predominately restricted to the context of secondary prevention and are often considered in isolation, not accounting for the substantial multimorbidity seen in those with hypertension.^[18]^

### POTENTIAL FOR PERSONALISED MEDICINE

Personalising therapy can promote the selection of the most effective drug for an individual, while avoiding therapies which are less effective for SBP reduction or associated with adverse effects. Substantial variability in the effectiveness of blood pressure treatment has been demonstrated in a systematic analysis of 135 population-based studies from 90 countries, underscoring the need for individualised approaches to hypertension management.^[2]^ Whilst contributed by differences in awareness and access to healthcare, there are also inter-individual and inter-population differences in treatment response and adherence^[19]^. Notably, differences in response between anti-hypertensive classes have been observed across 29 landmark hypertension trials.^[20]^ Randomised controlled trials (RCTs) confirm these major differences in blood pressure response and cardiovascular outcomes in underserved populations^[21–23]^, with a complex treatment effect that is also impacted by the lived environment.^[24]^ In The Precision Hypertension Care (PHYSIC) repeated cross-over trial of single antihypertensive drugs, substantial differences in treatment response were demonstrated, with a modelled gain of 4.4 mmHg for a personalised ‘best treatment’ approach versus a fixed choice.^[25]^ However, an evidence gap remains in guiding personalised therapy that frequently requires combination therapy^[26]^ in line with current hypertension guidelines.^[6,7]^ Metabolomics provides such an opportunity, quantifying thousands of metabolites that reflect an array of individual factors from genetic to environmental contributors that could be used to predict the response to treatment (pharmacometabolomics).^[27,28]^

The HYPERMARKER consortium was formed to address key evidence and implementation gaps – whether it is possible to personalise the choice of medication class for patients with hypertension, and how to empower patients to take an active role in shared treatment decisions. To enable personalised selection of antihypertensive drug classes, machine learning approaches will isolate and combine conventional clinical features with metabolomic profiles generated via state-of-the-art high-resolution mass spectrometry.

### THE HYPERMARKER TRIAL

The HYPERMARKER trial is the first RCT to evaluate a pharmacometabolomic-guided approach to aid antihypertensive drug class selection. The trial is a proof-of-concept, pragmatic, adaptive, open-label strategy trial embedded in routine clinical practice and will use stratified individual patient randomisation. The primary outcome is change in home SBP comparing the intervention and standard of care groups. A key objective of the HYPERMARKER trial is to develop, test and iterate a strategy of citizen engagement that can support personalised decision-making for hypertension treatment informing development of pharmacometabolomic-guided medical devices. The study is sponsored by the University of Birmingham and funded by EU Horizon and UK Research and Innovation.

## METHODS AND ANALYSIS

### PROTOCOL

The full protocol for the HYPERMARKER trial is available as a ***supplementary file***.

### PATIENT AND PUBLIC INVOLVEMENT

The HYPERMARKER project is supported by a patient and public involvement (PPI) group. PPI representatives were recruited through the European Heart Network, Brussels and represent each country taking part in the trial. The PPI group were involved in the study design and development of the trial protocol. Through focus groups, outcome measures were reviewed to ensure they represented the groups’ priorities and experience in prior treatment of hypertension. Trial procedures have been tested by the PPI team to ensure they are suitable, and any burden is balanced and acceptable, including the home blood pressure device, mobile application and patient reported outcome questionnaires. Participant recruitment material has been co-developed with the PPI team with participant information videos featuring members of the PPI team. The PPI groups will work with researchers during the trial with representation in the trial Joint Oversight Committee.

### ENROLMENT

Four hundred eligible patients will be invited to participate in the study during routine secondary care encounters across four hospital sites. The inclusion criteria are patients aged over 18 years old with a SBP ≥140 mmHg and a clinical indication for starting or adding antihypertensive therapy. Exclusion criteria are limited in order to maximise generalisability of findings ***(Table 1)***.

**Table 1.** Inclusion and Exclusion Criteria.

|  |  |
| --- | --- |
| Inclusion Criteria | • Age 18 years or older.<br>• SBP $\geq 140$ mmHg on any blood pressure recording method (office, home or ambulatory).<br>• Clinical indication for antihypertensive therapy. |
| Exclusion Criteria | • SBP $\geq 180$ mmHg on any blood pressure recording method (office, home or ambulatory).<br>• Three or more current anti-hypertensive medications.<br>• Potential secondary cause of hypertension, including but not limited to renovascular hypertension, endocrine conditions, chronic kidney disease, coarctation of the aorta, or medication-related.<br>• Planned intervention for hypertension, such as renal denervation.<br>• Severe kidney disease (eGFR $< 30$ mL/min).<br>• Diagnosis of known heart failure with a LVEF $< 40\%$.<br>• Stroke or myocardial infarction within the last 6 months.<br>• Pregnancy, planning for pregnancy, or breastfeeding.<br>• Participant whom the Clinical Investigator deems otherwise ineligible. |

SBP, systolic blood pressure; mmHg, millimetres of mercury; eGFR, estimated glomerular filtration rate; LVEF, left ventricular ejection fraction.

### INTERVENTION

The intervention (pharmacometabolomic approach) provides person-specific drug class decision support that clinicians may utilise when making their choice of blood pressure-lowering medication. This is generated using machine learning approaches that combine conventional clinical features with metabolomic profiles. Features of importance have been determined from pre-trial modelling across multiple pan-European cohorts (***Supplementary Table 1***). Standard of care involves usual clinical processes for choosing medication class and aligns with the 2024 European Society of Cardiology Guidelines for the Management of Elevated Blood Pressure and Hypertension.^[6]^

### RANDOMISATION & ALLOCATION

Following written consent, participants will be randomised in a 1:1 ratio to either initial standard of care (Group A) or initial use of the pharmacometabolomic approach for treatment selection (Group B). Randomisation will be stratified by site (four sites), participant age (18-69 & ≥70 years), and baseline office SBP (140-159mmHg or ≥160mmHg). As an open-label trial, both the investigator and participant will be aware of the allocation after randomisation. The pharmacometabolomic approach will be updated as additional data are obtained during the trial. Participants will have their medication class re-reviewed, with the updated pharmacometabolomic output provided to investigators for all participants, regardless of original randomised group, to ensure equity ***(Figure 2)***.

**Figure 2.**
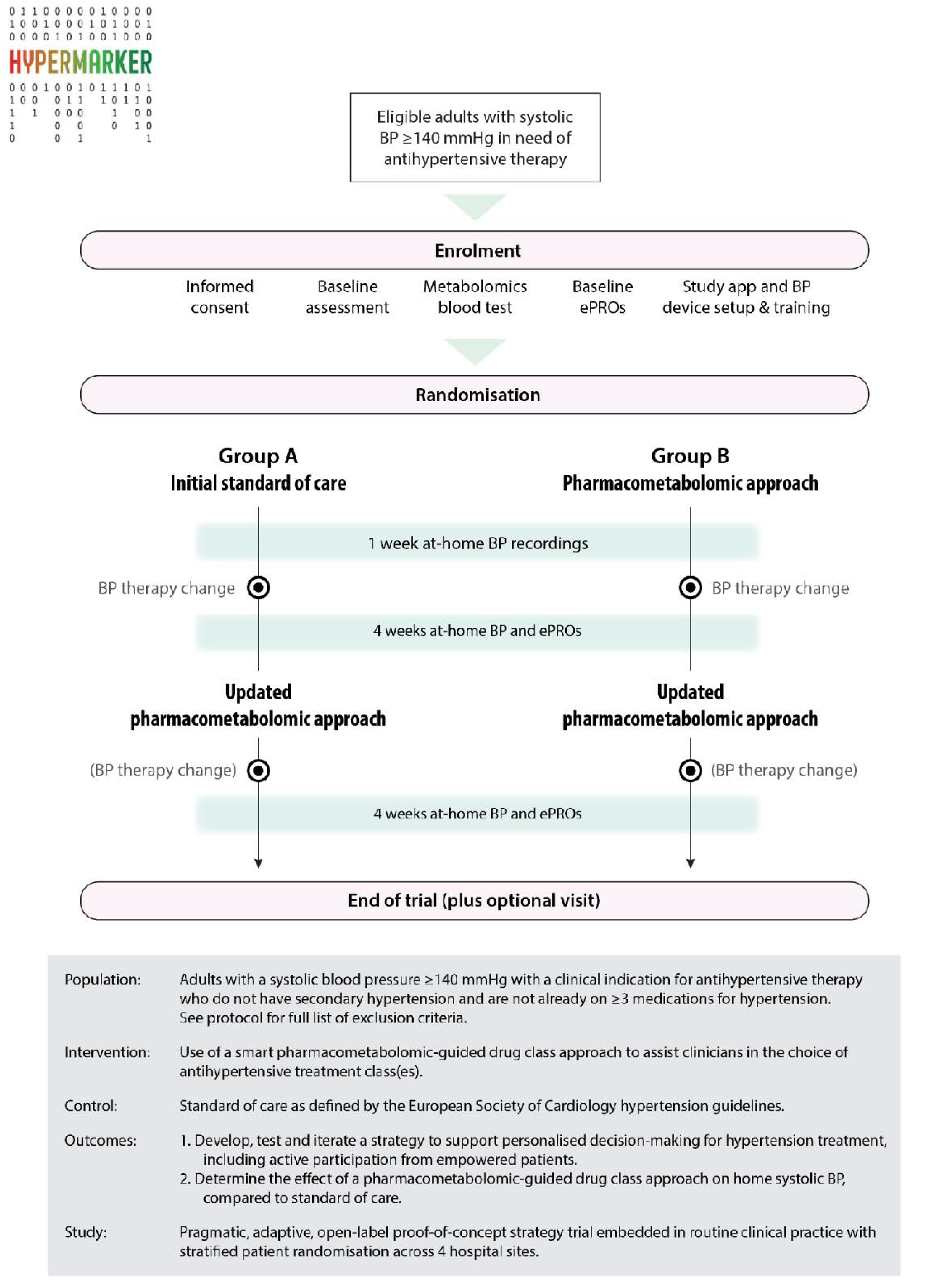
Schematic overview of HYPERMARKER trial design. *BP, blood pressure; ePROs, electronic patient-reported outcomes*.

### STUDY PROCEDURES

Baseline assessments at enrolment include collection of participant demographic data, medical and medication history, patient reported outcome questionnaires including quality of life and food intake questionnaires, and use of healthcare services in primary and secondary care. Individual metabolomic profiles are generated using a plasma sample taken at baseline and processed using a high-throughput liquid chromatography-mass spectrometry assay. Following enrolment, the trial will be conducted remotely with the provision of a home blood pressure device and smartphone application. Each participant will be asked to capture at least one week of home blood pressure measurements after randomisation and before changes to medications are made. A further 4 weeks of home blood pressure recordings will be requested after each medication change/review, with patient-reported quality of life, medication adherence, adverse events and healthcare consumption recorded at the end of each intervention phase via questionnaires completed using the participants own electronic device (***Figure 2***).

Home blood pressures will be measured using a semi-automatic, validated and CE-marked monitor (Rossmax Z5). Using a secure Bluetooth protocol, measurements are transferred to a mobile application (Viduet Health) on the participants phone and then securely transferred to a joint patient, physician & researcher portal to inform medication review. Home blood pressure measurements will be taken for at least 3 days per monitoring week, with participants asked to record at least two measurements, taken in the morning and evening on each day.

Plasma will be extracted from non-fasted EDTA samples collected at enrolment and frozen to −80 degrees Celsius prior to shipment to the Biomedical Metabolomics Facility Leiden in the Netherlands. Analysis will be performed via positive and negative ionisation liquid chromatography-mass spectrometry to identify both targeted and untargeted metabolites. The quantified metabolites of interest will be combined with conventional clinical variables within the machine learning-based tool to provide an estimated probability of beneficial treatment response according to hypertension drug class. Full details on the pharmacometabolomic algorithm will be published at the end of phase 2 of the trial to avoid contamination across randomised groups.

### STUDY OUTCOMES

The primary outcome is change in home SBP comparing the intervention and standard of care groups. The major secondary outcomes are rate of SBP change, change in diastolic blood pressure, proportion of participants achieving guideline defined target blood pressure, patient-reported adverse effects, quality of life, proportion of participants reporting treatment withdrawal and adherence. In addition, HYPERMARKER will demonstrate methods for direct patient involvement and empowerment, as well as healthcare pathways and health economic evaluation to underpin future implementation (***Supplementary Table 2*** for full list of trial outcomes).

### SAMPLE SIZE

The sample size of 400 patients using home SBP provides 99% power to detect a 1.5mmHg difference in SBP between the pharmacometabolomic approach and standard of care groups. Sample size calculations are based on the expected minimum of 12 baseline SBP measurements and 48 follow-up measurements by the end of the first phase, and assume a 2-sided alpha of 0.05, baseline SBP of 145 mmHg (SD 20), with correlation between SBP measurements of 0.7.^[29]^ Power is retained at >85% for a 2.5mmHg difference in home SBP even if only 3 baseline and 12 follow-up measurements are recorded (with correlation 0.5 and 2-sided alpha 0.05).

### STATISTICAL ANALYSIS

A statistical analysis plan will be drafted and agreed prior to database unlock. The primary analysis will follow the intention-to-treat principle, analysing all participants in their originally assigned groups regardless of compliance with the allocated group and with no imputation for any missing data. Sensitivity analyses will include ‘as-treated’ and ‘per-protocol’ analyses. The primary outcome of change in home SBP using multiple repeated measurements will be analysed using generalised linear models using a random-effects estimator and exchangeable correlation matrix. These account for all available SBP readings, accommodate variability in measurement frequency and intervals between participants, and account for correlation between repeated measures. Models will include adjustment for variables used in the randomised stratification (age and site), as well as gender, prior blood pressure treatment and relevant comorbidities. Secondary outcomes will be analysed using the same methods as described above (for continuous repeated outcomes), adjusted mean difference (for continuous outcomes), logistic or log-binomial regression (for categorical outcomes), chi-squared or Poisson regression (for count data), and incidence rate ratio (for adverse events). A p-value <0.05 will be considered statistically significant.

### TRIAL OVERSIGHT, MANAGEMENT AND REGISTRATION

Trial oversight, management and registration of the HYPERMARKER trial will be conducted in accordance with guidelines for Good Clinical Practice (GCP) and the Declaration of Helsinki. A joint oversight committee will comprise a Trial Steering Committee and an independent Data Monitoring Committee. Oversight will operate in accordance with trial-specific charters, based on the DAMOCLES recommendations.^[30]^ The trial is registered at ClinicalTrials.gov (NCT07294794) and ISRCTN (ISRCTN29385951). The trial protocol was developed in accordance with the Standard Protocol Items for Randomized Trials (SPIRIT) statement,^[31]^ the SPIRIT-PRO extension^[32]^ and the SPIRIT-Outcomes extension.^[33]^

### ETHICS AND DISSEMINATION

Ethical approval will be obtained for all trial sites. The North West – Greater Manchester West Research Ethics Committee (REC) (25/NW/0296) and the National Health Service Health Research Authority (IRAS project: 354889) have approved this study in England & Wales, UK. All findings of clinical relevance will be submitted to a suitable medical journal for publication after review by the oversight committees and the PPI team. The PPI team will provide a plain language summary of results that will be published alongside the scientific paper and sent to trial participants. Trial results will be reported in accordance with the CODE-EHR standards^[34]^ and CONSORT-AI reporting guidelines.^[35]^

## DISCUSSION

Personalising antihypertensive therapy selection to reflect individual variability in treatment response may help to address a key area of need and reduce the substantial and preventable burden of hypertension.^[25,36]^ The HYPERMARKER trial seeks to address key evidence gaps on how personalised therapy could be implemented, how patients can be empowered to take an active role in shared treatment decisions, and to determine whether individualised decisions on hypertension medication class are effective. HYPERMARKER will be the first time that person-specific drug class decisions using metabolomic profiles are tested within a randomised controlled trial, using recent advances in machine learning.

The pooled assessment of hypertension studies identifies a key need to individualise therapeutic approaches. Metabolomics offers a potential avenue to personalise antihypertensive therapy, with the composition and concentration of metabolites influenced by endogenous processes and their interactions with an individuals’ lived environment that reflects the multifaceted aetiology of hypertension ***(Figure 3)***. Metabolomics can facilitate differentiation of primary and endocrine hypertension, as well as identify potential markers of disease development and progression across different population groups.^[37–44]^ Potential metabolite biomarkers of blood pressure response to dietary and antihypertensive interventions have previously been demonstrated.^[45]^ N-methylglutamate, an amino acid linked to intake of fruits, was associated with a significant reduction in SBP response after 4 weeks of a low sodium diet compared with a high sodium control diet (p=0.031).^[46]^ Arachidonoyl-carnitine was associated with a reduced SBP response to atenolol by 3.15 mmHg (p = 0.006).^[47]^ Whilst the potential for pharmacometabolomics shows promise, most studies in this field only test the effect of monotherapies, which no longer aligns with current clinical guidelines. An evidence gap remains on how different dimensions of individual factors, including both clinical and metabolomic information can be integrated to guide personalised treatment selection.

**Figure 3.**
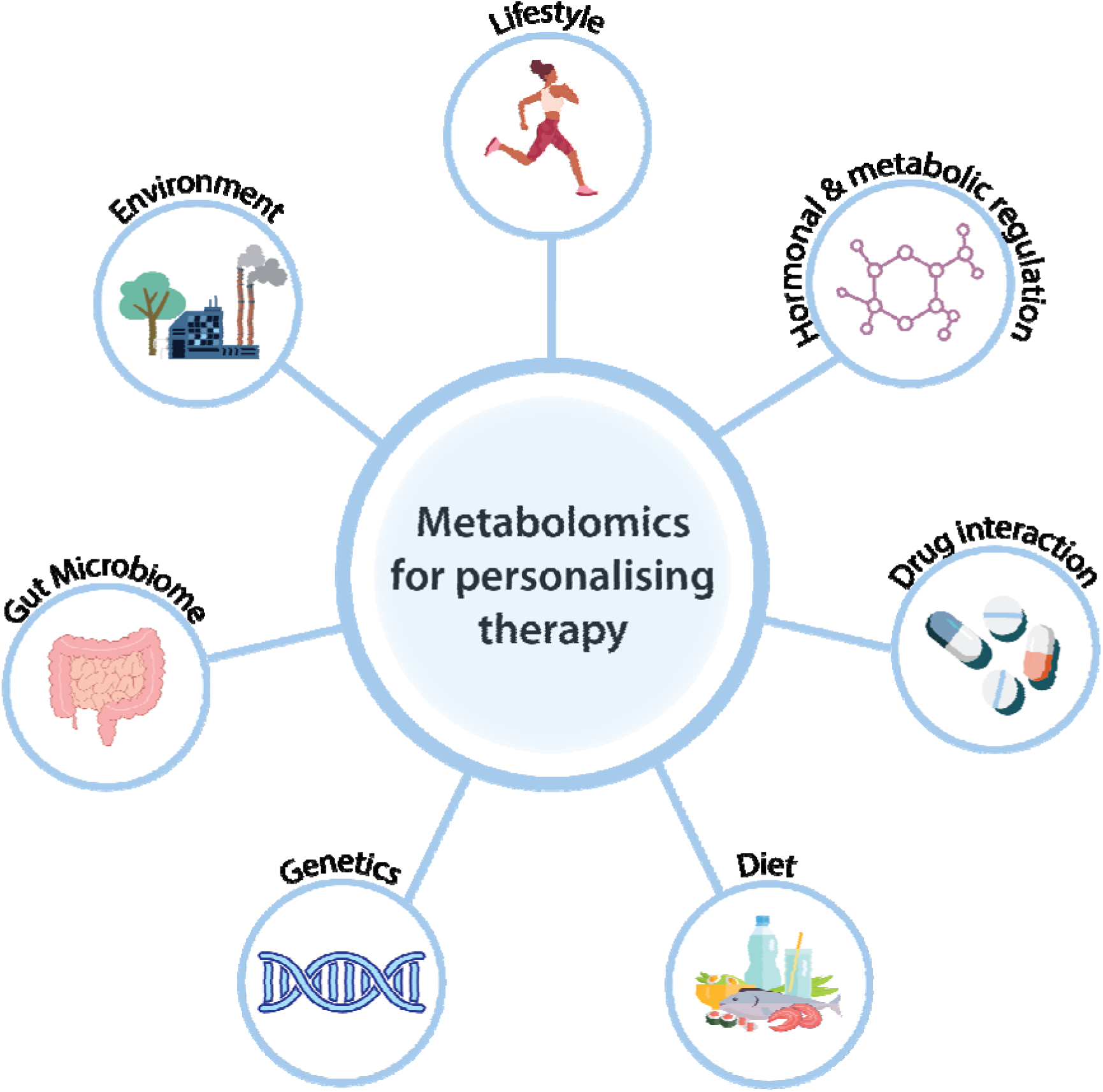
Span of individual patient factors related to metabolomics.

Machine learning can identify key variables from large, diverse biomedical and clinical datasets to generate highly accurate predictive models that can be validated across populations.^[48,49]^ It offers a potential pathway to personalised therapy and has already been applied to predict antihypertensive treatment response using standard clinical factors.^[50–52]^ However, large-scale studies that integrate metabolomic profiles into these models are lacking. Moreover, such models have not yet been tested in clinical trials limiting their potential for clinical implementation.^[53]^

Digital tools for personalised therapeutics can significantly improve compliance and empower patients to take an active role in their healthcare decisions.^[54,55]^ A combined trial intervention including patient self-management and monitoring, with tailored titration, lifestyle advice and behavioural support resulted in a mean difference in SBP of −3.4 mmHg (95% CI −6.1 to −0.8 mmHg) compared with usual care after 1 year (n=622).^[56]^ In an open=:Jlabel trial using daily home self=:Jmonitoring with linked smartphone app, personalised dosing of amlodipine achieved an SBP reduction of 11mmHg (95% CI 10–12, p<0.001, n=205) over 14=:Jweeks, with high patient-reported adherence (94%).^[57]^ However, an individual patient data meta-analysis of randomised trials found that self-monitoring alone does not improve BP, but requires a co-intervention.^[58]^

Hence the design of the HYPERMARKER trial, which includes PPI co-creation and digital tools that can link engaged patients with their healthcare provider to drive more efficient and cost-effective choices in blood pressure reduction. Use of out-of-office blood pressure assessment is implemented to enhance reliability and provide better correlation with end-organ damage and cardiovascular outcomes.^[59]^ However, as a proof-of-concept trial, the duration of HYPERMARKER is relatively short, and insufficient to assess cardiovascular outcomes. The four-week follow-up after each intervention aligns with the timing of SBP lowering effect from therapy^[60]^ as well as routine clinical practice.

## CONCLUSION

The HYPERMARKER proof-of-concept trial will evaluate personalised medication class selection for patients with hypertension using a pharmacometabolomic machine learning approach. The trial will determine whether empowered patients taking an active role in shared treatment decisions can result in more effective and efficient use of existing drug therapies, with fewer side effects and better adherence. The programme aims to address the current poor rates of blood pressure control, leading to prevention of hypertension-related morbidity and mortality, and improving the quality of life of individuals affected by one of the most prevalent chronic conditions worldwide.

## Supporting information

Supplemental File - Trial Protocol

## Data Availability

Data collection has not commenced.

## ACKNOWLEDGEMENTS

We would like to acknowledge the other members of the wider HYPERMARKER consortium and the significant contribution from the HYPERMARKER Patient and Public Involvement Team. We would also like to acknowledge our collaborators Kelly and Oscar at Viduet Health, Netherlands.

## CONTRIBUTORS

The manuscript was drafted by MC, DK and JK. MC and JK contributed equally to this paper. JK, AM, BL, MC and DK were the primary authors of the trial protocol and patient information. SZ, DE, AC, AWC and IB are members of the HYPERMARKER trial working group. FM-G, RS and WS are principal investigators. TH, DG and DK are members of the HYPERMARKER core leadership team and DK is the trial Chief Investigator. All authors contributed to the HYPERMARKER trial protocol or patient information and reviewed the manuscript for intellectual content.

