## Supplemental Tables for "Personalised approach to hypertension treatment: Rationale and design of the HYPERMARKER randomised trial"

**Supplementary information**

Supplementary Table 1. The HYPERMARKER cohorts

|  | **Atrial Fibrillation in High-Risk Individuals (AFHRI)** | **The Alpha Omega Cohort (AOC)** | **Utrecht Cardiovascular Cohorts – Second Manifestations of Arterial Disease (UCC-SMART)** | **Precision Hypertension Care (PHYSIC)** |
| --- | --- | --- | --- | --- |
| Population | Patients aged 18-85yrs referred to secondary care outpatient cardiology clinics | Aged 60-80 years old, with a verified history of myocardial infarction during the 10 years prior to study enrolment | Patients aged 18–90 years referred to secondary care for the management of cardiovascular disease or severe cardiovascular risk factors | Patients aged 40-75yrs, with mild hypertension 5-yrs prior to start of trial & untreated or on monotherapy at inclusion with a SBP of 140 and 179 mmHg at randomisation. |
| Study design | Prospective, single-centre cohort study | Multi-centre randomised double-blind placebo-controlled trial  Intervention: 400 mg/day EPA-DHA and/or 2 g/day ALA | Prospective single-centre cohort study | Single-centre, double-blind, randomised, repeated cross-over trial  Intervention: Four blood pressure-lowering drugs as monotherapy |
| Country | Germany | Netherlands | Netherlands | Sweden |
| Collection period for metabolomic samples used | 2017-2022 | 2002-2006 | 2008-2024 | 2017-2020 |
| Blood Pressure variables | Repeated office blood pressure | Repeated office blood pressure | Repeated office blood pressure | Repeated office and ambulatory blood pressure |

*EPA-DHA, Eicosapentaenoic Acid-Docosahexaenoic Acid; ALA, alphalinolenic acid*

Supplementary Table 2. Outcomes of the HYPERMARKER trial

Primary outcome

Change in home SBP will be derived from all available patient-measured SBP recordings in the study smartphone application, comparing the intervention and standard of care groups at the end of the first phase of the trial. This includes 1-week of monitoring after enrolment (anticipated minimum of 12 recordings) and at least 4-weeks of monitoring after therapy change (anticipated minimum of 48 recordings).

Secondary outcomes

The following secondary outcomes will compare the pharmacometabolomic-guided drug class approach and standard of care groups at the end of the first phase of the trial:

- Proportion of participants achieving a target home SBP of 120–129mmHg using the average of the final 3 days of blood pressure measurements.
- Proportion of participants reporting any treatment-related adverse effects compiled from the Summary of Product Characteristics from the different classes of anti-hypertensive medications.
- Proportion of participants reporting withdrawal of an anti-hypertensive medication.
- Proportion of participants reporting ≥90% adherence to prescribed anti-hypertensive medication.
- Rate of change in home SBP using all available SBP measurements, averaged per week.
- Change in home diastolic blood pressure derived from all available blood pressure recordings.

The following secondary outcomes will separately compare the original intervention, updated intervention and standard of care groups at the end of the second phase of the trial:

- Change in home SBP using all available SBP measurements, comparing the iterated pharmacometabolomic approach versus the initial pharmacometabolomic approach, and the iterated pharmacometabolomic approach versus initial standard of care.
- Patient-reported treatment-related side effects, comparing the iterated pharmacometabolomic approach versus the initial pharmacometabolomic approach, and the iterated pharmacometabolomic approach versus initial standard of care.

The following outcomes will apply across the phases of the trial comparing any pharmacometabolomic approach versus standard of care:

- Proportion and number of serious adverse events, including all-cause hospitalisation and death.
- Proportion and number of healthcare utilisation events, including details on hospitalisation (frequency, cause, type [outpatient, emergency, admission] and length of stay) and primary care interaction (frequency, cause and type [doctor, nurse, other allied health professional]).
- Patient-reported quality of life using the EQ-5D-5L summary index score and visual analogue scale.

Health Economics Outcomes

A separate health economics analysis will be conducted comparing any pharmacometabolomic approach versus standard of care to determine the net monetary benefit, incremental cost-effectiveness ratios, the cost-effectiveness planes and the cost-effectiveness acceptability curves, discounted at recommended values.

Exploratory Outcomes

- Change in metabolomic profile from baseline to (optional) follow-up blood sample, stratified by class of anti-hypertensive medication
- Association between dietary intake, metabolomic profile and blood pressure response to prescribed antihypertensive treatment

*SBP, systolic blood pressure; EQ-5D-5L, E*
