## Supplemental File - Trial Protocol for "Personalised approach to hypertension treatment: Rationale and design of the HYPERMARKER randomised trial"

**Personalised pharmacometabolomic-guided strategy trial to optimise treatment for hypertension (HYPERMARKER)**

**Protocol version number and date**

| | |
|---|---|
| **Protocol version number:** | **1.0** |
| **Protocol version date:** | **31-AUG-2025** |

**Research reference numbers**

| | |
|---|---|
| **IRAS number:** | **354889** |
| **Sponsor/RG number:** | **University of Birmingham, UK**<br>**RG_24-123** |
| **UK REC reference number:** | **North West – Greater Manchester West Research Ethics Committee (REC) (25/NW/0296)** |
| **Public registry number:** | **Clinicaltrials.gov: NCT07294794**<br>**ISRCTN: ISRCTN29385951** |
| **Funder number:** | **Horizon Europe 101095480; UKRI 10061996** |

#### Signature page

The undersigned confirm that the following protocol has been agreed and accepted and that the Chief Investigator (CI) agrees to adhere to the signed University of Birmingham's sponsorship CI declaration.

I agree to ensure that the confidential information contained in this document will not be used for any other purpose other than the evaluation or conduct of the investigation without the prior written consent of the Sponsor.

I also confirm that I will make the findings publicly available through publication or other dissemination tools without any unnecessary delay and that an honest accurate and transparent account of the project will be given; and that any discrepancies from the project as planned in this protocol will be explained.

| | |
|---|---|
| **Full project title:** | **Personalised pharmacometabolomic-guided strategy trial to optimise treatment for hypertension (HYPERMARKER)** |
| **Protocol version number:** | |
| **Protocol version date:** | |

| **Chief Investigator (CI)** |
|---|
| **Name:** |
| **Date:** |
| **Signature:** |

Sponsor statement
Where the University of Birmingham takes on the sponsor role for protocol development oversight, the signing of the IRAS form by the sponsor will serve as confirmation of approval of this protocol.

### PRINCIPAL INVESTIGATOR SIGNATURE PAGE

I have read and agree to the protocol, as detailed in this document. I agree to adhere to the protocol as outlined and agree that any suggested changes to the protocol must be approved by the Trial Steering Committee prior to seeking approval from the Research Ethics Committee and Regulatory Authority.

I am aware of my responsibilities as an Investigator under the guidelines of Good Clinical Practice (GCP), the Declaration of Helsinki, local regulations (as applicable) and the trial protocol and I agree to conduct the trial according to these guidelines and to appropriately direct and assist the staff under my control who will be involved in the trial.

**Site Principal Investigator:**

Name:

...............................................................................

Signature: Signature Date:

............................................................................... ....../....../......

Site Name:

.........................................................................................................................................

Protocol Date: ....../....../......

Protocol Version: …………………

#### KEY STUDY CONTACTS

| Role | Contact |
| --- | --- |
| **Trial Management Group** | **Core team:**<br><br>Diederick Grobbee, Professor of Clinical Epidemiology, University Medical Centre Utrecht & Julius Center for Health Sciences and Primary Care, Netherlands:, +31 621162958<br><br>Jonathan Knikman, Assistant Professor, Julius Center for Health Sciences and Primary Care, Netherlands,, tel: +31 629686273<br><br>Alastair Mobley, Trial Team Manager, University of Birmingham, Birmingham, UK:, +44 1213718145<br><br>Dipak Kotecha, Professor of Cardiology, Institute of Cardiovascular Sciences, University of Birmingham & University Hospitals Birmingham NHS Foundation Trust (**Chief Investigator**):; +44 1213718122<br><br>**Site Principal Investigators:**<br><br>Matthew Chapman, Clinical Research Fellow, University Hospitals Birmingham NHS Foundation Trust, Birmingham, UK:, +44 1213718145<br><br>Wilko Spiering, Associate Professor, University Medical Centre Utrecht, Netherlands, Department of Vascular Medicine,, tel: +31 887571188<br><br>Fernando Martinez-Garcia, Internal Medicine Consultant, INCLIVA, Biomedical Research Institute, Valencia, Spain:, +34 679663209<br><br>Renate Schnabel, Professor in Cardiology, University Heart and Vascular Center Hamburg, University Medical Center Hamburg-Eppendorf, Hamburg, Germany:, +49 40 741053979 |
| **Trial Joint Oversight Committee** | Chair of the Independent Trial Steering Committee: Prof Bryan Williams, Chair of Medicine, University College London and Chief Scientific & Medical Officer, British Heart Foundation, London, UK:, +44 2076792000<br><br>Chair of the Independent Data Management & Safety Committee: Prof. Neil Poulter, Professor of Preventive Cardiovascular Medicine, Imperial College London, London, UK:, +44 2075943446<br><br>Other members of the Joint Oversight Committee:<br><br>Pat Mooney, Patient & Public Involvement Representative<br><br>Prof. Tine de Backer, Professor of Cardiology, Ghent University, Ghent, Belgium:, +32 93323459<br><br>Dr Peter Van de Ven, Associate Professor of Clinical Trial Methodology and Statistics, Department of Data Science and Biostatistics, Julius Center, University Medical Centre Utrecht, Netherlands,, +31 887555555 |

| Role | Contact |
|---|---|
| **Health Economics Team** | Alex Carter, Associate Professor, London School of Economics and Political Science, London, UK:, +44 2079556959<br>Seraphine Zeitouny, Research Fellow, London School of Economics and Political Science, London, UK:, +44 2079556959 |

#### TRIAL SYNOPSIS

| | |
|---|---|
| Title | Personalised pharmacometabolomic-guided strategy trial to optimise treatment for hypertension (HYPERMARKER) |
| Acronym | HYPERMARKER |
| Funder | Horizon Europe and UKRI |
| Sponsor | University of Birmingham |
| Trial sites | INCLIVA Instituto de Investigación Sanitaria, Spain; University Hospitals Birmingham NHS Foundation Trust, United Kingdom; University Medical Center Hamburg-Eppendorf, Germany; University Medical Center Utrecht, the Netherlands |
| Trial design | Pragmatic, adaptive, proof-of-concept, open-label strategy trial embedded in routine clinical practice with individual patient randomisation |
| Population | Patients in routine clinical practice in need of antihypertensive therapy |
| Trial intervention | Use of a smart pharmacometabolomic approach to assist clinicians in the choice of antihypertensive treatment class(es) versus usual standard of care |
| Primary objectives | Develop, test and iterate a strategy to support personalised decision-making for hypertension treatment, including active participation from empowered patients<br><br>Determine the effect of a pharmacometabolomic-guided drug class approach on home systolic blood pressure (SBP), compared to standard of care (null hypothesis: no difference in home SBP) |
| Secondary objectives | • Proportion of participants achieving a target home SBP of 120–129mmHg<br>• Patient-reported treatment-related side effects<br>• Patient-reported treatment withdrawal<br>• Patient-reported treatment adherence<br>• Rate of change in home SBP<br>• Change in home diastolic blood pressure<br>• Change in home SBP with an iterated pharmacometabolomic approach<br>• Patient-reported treatment-related side effects with an iterated pharmacometabolomic approach<br>• Number of serious adverse events<br>• Number of hospital and primary care visits<br>• Patient-reported quality of life<br>• Change in metabolomic profile (exploratory)<br>• Association with dietary intake (exploratory) |
| Health economic outcomes | • Life years and quality-adjusted life year (QALY) gained<br>• Net monetary benefits of the pharmacometabolomic approach<br>• Difference in healthcare utilisation and cost estimates between standard care and the pharmacometabolomic approach |
| Monitoring duration | 1 week of home blood pressure monitoring at baseline, 4 weeks following initial therapy and 4 weeks following iteration of therapy (total 9-16 weeks) |
| Estimated number of participants | 400 participants |
| Inclusion criteria | 1. SBP ≥140 mmHg<br>2. Age 18 years or older |

| | |
|---|---|
| | 3. Clinical indication for antihypertensive therapy |
| Exclusion criteria | 1. SBP ≥180 mmHg<br>2. Potential secondary cause of hypertension, including but not limited to renovascular hypertension, endocrine conditions, chronic kidney disease, coarctation of the aorta or medication related<br>3. Three or more current anti-hypertensive medications<br>4. Planned intervention for hypertension, such as renal denervation<br>5. Severe kidney disease (estimated glomerular filtration rate <30 mL/min)<br>6. Diagnosis of known heart failure with left ventricular ejection fraction <40%<br>7. Stroke or myocardial infarction within the last 6 months<br>8. Pregnancy, planning for pregnancy, or breastfeeding<br>9. Participant whom the Clinical Investigator deems otherwise ineligible |

#### FUNDING

This study is funded by an EU Horizon funding grant (European Health and Digital Executive Agency (HADEA), European Commission, B-1049, Brussels, Belgium) with additional funding via the UK Government's Horizon Europe Guarantee from UK Research and Innovation Innovate UK (Polaris House, North Star Avenue, Swindon, SN2 1FL, United Kingdom).

#### ROLES & RESPONSIBILITIES OF MANAGEMENT COMMITTEES

A joint oversight committee comprising a Trial Steering Committee (TSC) and Data Monitoring Committee (DMC) will be engaged for this trial, their role is outlined in section 9.4 of this document.

The HYPERMARKER project is supported by a patient and public involvement (PPI) group which consists of members from each country from which the trial will recruit (the United Kingdom, Spain, Netherlands & Germany). The PPI group have been involved in the trial design and content of this protocol. The PPI group will work with researchers during the trial with representation in the trial steering committee.

#### PROTOCOL CONTRIBUTORS

This protocol has been written by Alastair Mobley, Alex Carter, Bart Lagerwaard, Diederick Grobbee, Dipak Kotecha, Jonathan Knikman, Matthew Chapman, and Seraphine Zeitouny.

The sponsor has reviewed the ethical and regulatory aspects of this protocol but has not been invovled in the project design. They will not be involved in the data analysis and interpretation, manuscript writing, nor dissemination of results.

The funder has not been involved in the project design, nor will they be involved in the conduct, data analysis and interpretation, manuscript writing or dissemination of results.

### TRIAL FLOWCHART

*Figure 1: Schematic overview of trial design*

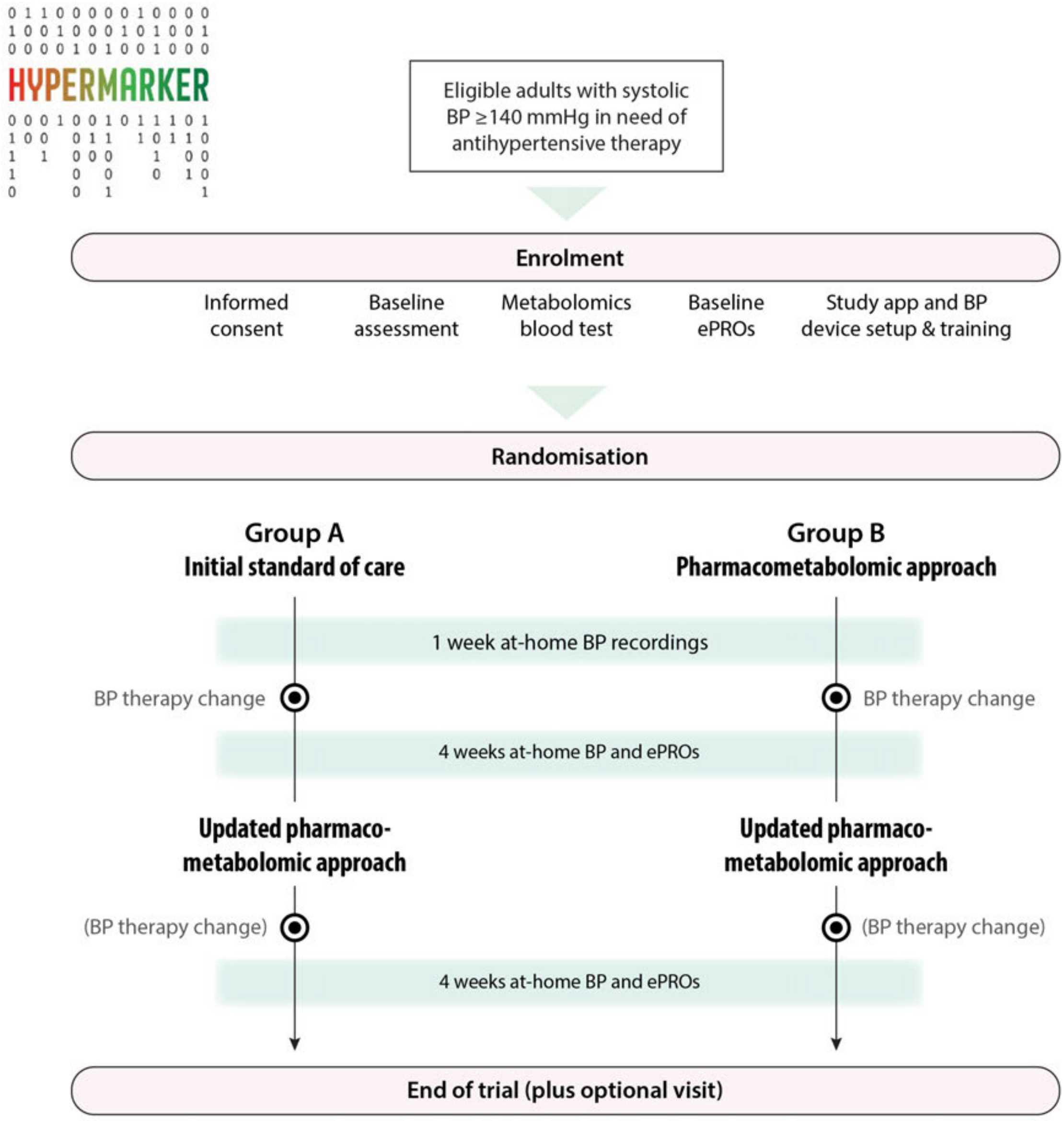

| | |
|---|---|
| Population: | Adults with a systolic blood pressure ≥140 mmHg with a clinical indication for antihypertensive therapy who do not have secondary hypertension and are not already on ≥3 medications for hypertension. See protocol for full list of exclusion criteria. |
| Intervention: | Use of a smart pharmacometabolomic-guided drug class approach to assist clinicians in the choice of antihypertensive treatment class(es). |
| Control: | Standard of care as defined by the European Society of Cardiology hypertension guidelines. |
| Outcomes: | 1. Develop, test and iterate a strategy to support personalised decision-making for hypertension treatment, including active participation from empowered patients. 2. Determine the effect of a pharmacometabolomic-guided drug class approach on home systolic BP, compared to standard of care. |
| Study: | Pragmatic, adaptive, open-label proof-of-concept strategy trial embedded in routine clinical practice with stratified patient randomisation across 4 hospital sites in Spain, Germany, UK & the Netherlands. |

BP, blood pressure; ePROs, electronic patient-reported outcomes.

#### ABBREVIATIONS

| | |
|---|---|
| AE | Adverse event |
| AI | Artificial Intelligence |
| AR | Adverse Reaction |
| BP | Blood Pressure |
| CI | Chief Investigator |
| DBP | Diastolic Blood Pressure |
| DMC | Data Management Committee |
| eCRF | Electronic Case Report Form |
| ePROs | Electronic Patient-Reported Outcomes |
| ESC | European Society of Cardiology |
| EU | European Union |
| GCP | Good Clinical Practice |
| GDPR | General Data Protection Regulation |
| ICF | Informed Consent Form |
| ISF | Investigator Site File |
| PIL | Patient Information Leaflet |
| PPI | Patient and Public Involvement |
| REC | Research Ethics Committee |
| REDCap | Research Electronic Data Capture |
| SAE | Serious Adverse Event |
| SAP | Statistical Analysis Plan |
| SBP | Systolic Blood Pressure |
| TMG | Trial Management Group |
| TSC | Trial Steering Committee |
| WOCBP | Women of Childbearing Age |

### PLAIN ENGLISH SUMMARY

High blood pressure (hypertension) affects around 1 in 3 adults and can cause serious health problems such as a stroke or heart attack. Medication to lower blood pressure is proven to reduce the risk of these complications. There are many different drugs that can treat high blood pressure, and a combination of tablets at lower dose is usually the best option. However, choosing the right combination for each person is difficult, which can lead to side effects or poor control of blood pressure.

HYPERMARKER is testing whether providing doctors with additional information when they decide on which medication to prescribe can improve a patient's overall blood pressure management. This includes relevant clinical information and personalised results from blood tests, brought together using computer programs (machine learning). The blood tests check for small substances naturally produced by the body called metabolites that may indicate how a patient might react to certain medications; this is termed a pharmacometabolomic approach.

This clinical trial has been developed with the support of a Patient and Public Involvement Team. We plan to recruit four hundred people across four sites in the UK, Spain, Netherlands & Germany. To be included in the study requires a recent high blood pressure reading that needs treatment. All participants will need to provide written informed consent to join the trial. They will have a blood sample taken for the metabolite test, and be provided with a blood pressure machine. The blood pressure machine connects to a smartphone application to keep a record of their measurements to allow the trial to be performed at home. The main outcome of the trial is home systolic blood pressure recordings, checked regularly for 1 week before and 4 weeks after each medication review. Participants will also be asked to complete questionnaires using their electronic device (smartphone, tablet or computer) to assess their views on their own health, how they are finding the treatment, and their use of healthcare services.

Each patient will be assigned randomly to one of two groups (A or B) so that we can test and improve how the pharmacometabolomic approach works. Everyone will have their medications reviewed and if needed, changed by their treating doctor. Those in Group A will first receive medication as per normal routine practice (standard of care; without any additional information). Those in Group B will have medication prescribed by a doctor with access to the pharmacometabolomic information up-front. In both groups, the healthcare teams will later receive updated information from the pharmacometabolomic approach and decide whether any changes are needed, so that all participants receive informed blood pressure management over the course of the research.

At the end of the trial, which will last 9-16 weeks, all participants and their usual doctor (such as general practitioner or hospital doctor) will be given a summary of their blood pressure readings, prescribed medications and response, so they can decide on longer-term blood pressure treatment.

### 1 INTRODUCTION AND RATIONALE

#### 1.1 Burden of hypertension

Hypertension is defined as a systolic blood pressure (SBP) ≥140 mmHg and/or diastolic blood pressure (DBP) ≥90 mmHg by the European Society of Cardiology (ESC).[1] Hypertension is common, with over 30% of adults estimated to be affected worldwide (approximately 1.3 billion).[2] Hypertension is the most important modifiable risk factor for all-cause and cardiovascular morbidity and mortality globally.[3] Systolic hypertension leads to a broad variety of diseases with considerable impact on both patients and healthcare systems, ranging from all forms of heart disease (in particular, coronary disease, heart failure, and atrial fibrillation), to strokes, kidney disease, and vascular dementia. These major public health concerns are potentially preventable with effective blood pressure treatment.[4]

The global burden of hypertension is increasing. Prevalence doubled between 1990-2019, with greatest prevalence in low- and middle-income countries.[4] It is estimated that 46% of adults are unaware they have hypertension until opportunistic identification occurs, or the person suffers an adverse medical event. Even with diagnosis, only a minority of patients achieve their target blood pressure control.[4] This is critical, as each 5-mmHg reduction of systolic blood pressure can reduce the risk of major cardiovascular events by around 10%.[5,6]

The overall economic costs are equally staggering, corresponding to 10% of EU healthcare expenditure.[7] The management of hypertension is estimated to account for 10% of primary care visits.[8] These substantial hypertension-related health and economic burdens make dealing with hypertension an immediate public health priority.

#### 1.2 Need for personalised treatment

While effective drugs are available for hypertension, overall control of blood pressure is poor. In a cross-sectional study of 153,996 adults aged 35 to 70 years (from 628 global communities), only 33% of those receiving treatment had adequate blood pressure control.[9] In the European Study on Cardiovascular Risk Prevention and Management in Usual Daily Practice, less than 40% of hypertensive patients achieved target blood pressure.[10] The World Health Organisation (WHO) has repeatedly stressed the urgency of improving blood pressure control in patients and populations, and considers this one of its key global targets.[11]

Although lifestyle measures may initially be trialled in hypertension management, most patients require drug therapy. Common strategies to improve blood pressure control are focused on the addition or substitution of drugs, but these approaches are rarely scientifically based. Most clinicians will choose therapies they are familiar with, rather than according to individual patient characteristics. This strategy increases the risk of medication-related adverse effects that can lead to poor adherence.[12,13] One year after initiation, adherence to blood pressure medication is less than 50%.[14] The conventional approach to treatment selection also ignores the potential for an individual's differential response to therapy. The underlying reason for poor blood pressure control is often overlooked, for example related to interactions with lifestyle, other medications, and the increasing problem of multi-morbidity. In a UK community cohort of 295,487 people first diagnosed with hypertension, 35% had two or more comorbidities.[15]

The effectiveness of drug treatment can be influenced by several factors including genetics, the environment, the diet of the patient (especially salt intake), and the gut microbiome.[16] Personalised treatment choice in clinical care has attracted considerable interest, for example using genomic data for adaptive treatment regimes for cancer.[17] While several studies support an important contribution of genetic polymorphisms on blood pressure response to antihypertensive therapy, studies have failed to detect significant effects or replicate previous findings.[18] These inconsistencies may be related to differences in the distributions of polymorphisms, or to epigenomic alterations that could mask the contribution of variants.[19] Downstream measures of the interactions between genes and the environment, such as an individual's metabolomic profile, could be a better candidate to personalise treatment choice for high blood pressure.

##### 1.3 Pharmacometabolomic optimised hypertension treatment

A persons metabolomic profile comprises endogenous metabolites produced by the body, such as amino acids, sugars, organic acids, lipids, vitamins, growth factors, nucleosides and nucleotides.[20] The composition and concentration of endogenous metabolites is influenced by a variety of factors.[20] An advantage of metabolomics compared to genomics is that it provides a direct readout of the current metabolic state of an individual. The metabolomic profile can inform about health state, combining multiple stacked layers of an individual's physiology to predict treatment response in that person.[21] By including multiple aspects in context of their causal and temporal interactions, a personalised treatment approach for hypertension could be achieved, in which inter-individual variability is considered.[19] This approach can also deal with retention of anti-hypertensive effect, in particular to enhance the low rates of adherence to long-term therapy, by reducing occurrence of medication-related adverse effects.[22] Digital tools for personalised therapeutics can significantly improve compliance.[23] There is also the potential for digital platforms to empower patients to take an active role in their healthcare decisions.[24]

##### 1.4 HYPERMARKER summary

HYPERMARKER seeks to use individual patient metabolomic profiles to assist clinicians in their choice of antihypertensive drug class(es), in line with current clinical practice guideline recommendations. This protocol outlines the design of the HYPERMARKER trial, a proof-of-concept pragmatic strategy trial co-developed with Patient & Public Involvement (PPI) to investigate how pharmacometabolomics can be integrated into routine practice to personalise routine treatment of hypertension.

#### 2 OBJECTIVES

##### 2.1 Primary Objectives

- Develop, test and iterate a strategy to support personalised decision-making for hypertension treatment, including active participation from empowered patients.
- Determine the effect of a pharmacometabolomic-guided drug class approach on home SBP, compared to standard of care (null hypothesis: no difference in home SBP).

##### 2.2 Secondary objectives

- Compare the proportion of participants achieving a target home SBP of 120–129mmHg with a pharmacometabolomic approach versus standard of care.
- Compare the incidence of treatment-related adverse effects with a pharmacometabolomic approach versus standard of care.
- Compare treatment withdrawal due to patient-reported adverse events or adverse reactions with a pharmacometabolomic approach versus standard of care.
- Compare patient-reported adherence to antihypertensive treatment with a pharmacometabolomic approach versus standard of care.
- Evaluate the rate of change in home SBP following new therapy, comparing a pharmacometabolomic approach with standard of care.
- Compare home diastolic blood pressure with a pharmacometabolomic approach versus standard of care.
- Determine the effect of an iterated and updated pharmacometabolomic approach on home SBP.
- Determine the effect of an iterated and updated pharmacometabolomic approach on treatment-related adverse effects.
- Compare the incidence of serious adverse events with any pharmacometabolomic approach versus standard of care.
- Compare the incidence of healthcare utilisation events with any pharmacometabolomic approach versus standard of care.
- Identify changes in patient-reported quality of life across the different treatment phases of the trial.
- Explore changes in metabolomic profile after hypertension treatment, stratified by drug class.

- Explore how dietary intake impacts on metabolomic profiles and blood pressure response to antihypertensive treatment.

##### 2.3 Health economic objectives

- Conduct a cost-utility analysis using cost and quality of life (EQ-5D-5L) to derive a cost per Quality-Adjusted Life year (QALY) gained comparing the pharmacometabolomic approach with standard of care.
- Conduct a benefit analysis expressing the net monetary benefits/costs of the intervention in the trial setting comparing the pharmacometabolomic approach with standard of care.
- Develop a probabilistic decision analytical model comprising of (1) a decision tree that captures the short-term clinical outcomes and costs associated with the two arms of the RCT in phase one, with a time horizon defined by the duration of the trial follow-up (12 weeks).
- Using process mapping methods, map clinical sequelae and resource utilisation across various pathways of care in the European healthcare setting.
- Conduct time-driven activity-based costing across pathway models and identify and rank cost drivers.

#### 3 STUDY PLAN AND DESIGN

##### 3.1 Trial design and setting

HYPERMARKER is a proof-of-concept, pragmatic, adaptive, open-label strategy trial embedded in routine clinical practice with stratified individual patient randomisation. The setting is secondary care, including four hospital sites in four countries (Germany, the Netherlands, Spain and the United Kingdom). Except for the enrolment process and optional final blood test, the trial can be managed remotely, or with in-person visits as preferred by sites and participants.

The intervention will combine metabolomic and clinical data using machine learning to provide additional information clinical investigators may utilise in their choice of blood pressure-lowering medication class for individual patients (pharmacometabolomic approach). ***Figure 1*** provides a schematic overview, and further detail on the intervention is provided in Section 5. The trial is organised into two phases to iterate and improve the pharmacometabolomic approach, and ensure that all patients included in the trial have access to the intervention. In the first phase, participants will be randomised to usual standard of care (group A) for treatment selection, or initial pharmacometabolomic approach (group B). In the second phase, participants originally randomised to group A will have their medications re-reviewed by the clinical investigator with access to the latest iteration of the pharmacometabolomic approach. Similarly, those originally randomised to group B will also potentially benefit from updates to the pharmacometabolomic approach during the course of the trial.

##### 3.2 Enrolment

HYPERMARKER is a pragmatic clinical trial, and patients will be recruited by opportunity within the recruitment period for each site. After written informed consent, the participant will be randomised and a blood sample collected to assess their metabolomic profile. Patient demographics, medical and medication history, comorbidity burden and patient-reported outcomes are collected. The study team will set up the home blood pressure monitoring device, demonstrate to the participant, and connect it to their own mobile device. Each participant will be asked to capture at least one week of home blood pressure measurements following randomisation. Patients, clinicians and study researchers will be able to access blood pressure values and trends using the secure online trial portal.

#### 3.3 Randomisation and treatment allocation

Timing: Participants can only be randomised after all eligibility criteria have been confirmed and informed consent is documented.

Process: Randomisation will be stratified by site (four sites), participant age (18-69 & ≥70 years), and baseline SBP (SBP 140-159mmHg or ≥160mmHg), allocating the participants 1:1 to either Group A (initial standard of care) or Group B (up-front pharmacometabolomic approach). Investigators will randomise participants directly through the online trial platform using the Research Electronic Data Capture (REDCap) system. In the event that the trial platform is unavailable or has technical issues, randomisation will be delayed (no backup paper system is planned to minimise logistical burden for sites).

Concealment of allocation: The Investigator is blinded to the allocation sequence. As an open-label trial, both the Investigator and participant will be aware of the allocation after randomisation, with the Investigator responsible for informing the participant about the randomised allocation.

Procedures for handling incorrectly enrolled or randomised patients: Patients who fail to meet the eligibility criteria should not, under any circumstances, be enrolled. Patients who are enrolled, but subsequently found not to meet all the eligibility criteria must not be randomised and must be withdrawn from the trial. Where a patient does not meet all the eligibility criteria but is randomised in error, the Investigator should inform the Sponsor (or delegate) via the Trial Coordinator immediately, and a discussion should occur between the Sponsor (or delegate) and the Investigator regarding whether to continue or discontinue participation. The Sponsor (or delegate) must ensure all decisions are appropriately documented.

Procedure where a participant withdraws from the trial: If a participant withdraws then their randomisation code cannot be reused. If a participant withdraws, they can be replaced.

#### 3.4 Group A

Any changes or additional medication(s) will be initiated by the Clinical Investigator after the participant has completed at least one week of home blood pressure measurements. Choice of therapy will be left to the Investigator based on their usual clinical approach, and without the additional pharmacometabolomic information. Four weeks of home blood pressure monitoring will be requested of each participant (with reminders sent via patient smartphone application or text message as needed). In the second phase, investigators are provided with the output from the latest iteration of the pharmacometabolomic approach when making their own clinical decision on what medication to prescribe. Clinical staff are encouraged to prescribe appropriate therapy, ideally aligning with the pharmacometabolomic drug class approach if deemed clinically appropriate. Participants will be informed of their updated clinician determined therapy suggestions/changes via a message on the smartphone application (sent by the investigator or delegate) unless an alternative method (text message, email or phone call) has been agreed with the participant and site. The participant is then requested to complete a further 4-weeks of home blood pressure monitoring via the smartphone application regardless of any therapy change.

#### 3.5 Group B

The output from the pharmacometabolomic approach will be communicated within a 4-6 week period to investigators. Clinical staff will then be expected to prescribe therapy of their own choosing, ideally aligning with the pharmacometabolomic drug class approach if deemed clinically appropriate. Participants will receive notification of the clinician suggested/prescribed therapy via a message on the smartphone application (sent by the investigator or delegate) or alternative agreed method (text message, email or phone call). Four weeks of home blood pressure monitoring will be requested of each participant following the start or change to any therapy (with reminders sent via the patient smartphone application or text message as needed). In the second phase, Clinical Investigators will receive an update from the latest iteration of the pharmacometabolomic approach and may choose to change the prescribed medication if clinically appropriate. Changes will be disseminated via the smartphone application messages (sent by the research team) or alternative agreed method (text message, email or phone call), with a further 4-weeks of home blood pressure monitoring requested regardless of any therapy change.

##### 3.6 End of trial

Patient-reported outcome questionnaires will be completed remotely using any electronic device/system (Phone, tablet, laptop desktop PC) via the REDCap system web-browser. The participants blood pressure averages across the duration of the trial along with prescribed therapy will be provided to Investigators, their usual primary healthcare provider and participants in PDF format to help advise future clinical management. Each participant will be asked to attend their local research site to have an optional blood sample to assess changes in their metabolomic profile; this is for research purposes only and will not contribute to clinical care.

#### 4 STUDY POPULATION

##### 4.1 Population

HYPERMARKER will recruit at least 400 patients who fulfil the trial selection criteria. Eligibility should be confirmed by medically-qualified personnel. Potentially eligible participants can be identified from referrals, hospital clinics, ambulatory care centres, during hospital admissions or via direct outreach to patients using advertisements.

##### 4.2 Inclusion criteria

1. Age 18 years or older.
2. SBP ≥140 mmHg on any blood pressure recording method (office, home or ambulatory).
3. Clinical indication for antihypertensive therapy.

##### 4.3 Exclusion criteria

1. SBP ≥180 mmHg on any blood pressure recording method (office, home or ambulatory).
2. Three or more current anti-hypertensive medications.
3. Potential secondary cause of hypertension, including but not limited to renovascular hypertension, endocrine conditions, chronic kidney disease, coarctation of the aorta, or medication-related.
4. Planned intervention for hypertension, such as renal denervation.
5. Severe kidney disease (estimated glomerular filtration rate <30 mL/min).
6. Diagnosis of known heart failure with left ventricular ejection fraction <40%.
7. Stroke or myocardial infarction within the last 6 months.
8. Pregnancy, planning for pregnancy, or breastfeeding.
9. Participant whom the Clinical Investigator deems otherwise ineligible.

##### 4.4 Screen failures

A screen failure occurs when a participant who consents to participate in the clinical study does not subsequently start antihypertensive treatment. A minimal set of screen failure information is required to ensure transparent reporting of screen failure participants to meet the Consolidated Standard of Reporting Trials (CONSORT) publishing requirements and to respond to queries from regulatory authorities. Minimal information includes demography, screen failure details, eligibility criteria, and any serious adverse events (SAEs). Participants who do not meet the criteria for participation may be rescreened in case the original screen failure was due to reasons expected to change at rescreening. Rescreened participants will be assigned new pre-screening/participant numbers.

#### 4.5 Informed consent process

It will be the responsibility of the Investigator to obtain written informed consent for each participant prior to performing any trial related procedure. If local practice allows, this responsibility may be delegated by the Principal Investigator to a Research Nurse or clinician as captured on the Site Signature and Delegation Log. Investigators or delegate(s) will ensure that they adequately explain the aim, trial intervention, anticipated benefits and potential hazards of taking part in the trial to the participant. They will also stress that participation is voluntary and that the participant is free to withdraw from the trial at any time. Participants will be given the opportunity to ask questions and a Participant Information Leaflet (PIL) will be provided. The PIL contains a link to a Participant Information Video (PIV) developed with and featuring the PPI team. This study augments usual care and, by current guidelines, patients that meet the selection criteria require a change in their medical therapy. To ensure this clinical need is met, participants may be immediately entered into the study. They can subsequently withdraw without clinical consequence if after further consideration of the PIL and PIV they do not feel the trial meets their needs. Where the participant lacks capacity for consent, they should not be recruited.

If the participant expresses an interest in participating in the trial they will be asked to sign and date the latest version of the Informed Consent Form (ICF). The ICF can be a written form, or a remote electronic consent form, depending on usual practice and legal requirements at each site. The participant must give explicit consent for regulatory authorities, members of the research team and representatives of the sponsor to be given access to trial records. The ICF will include consent to communicate with the participant's usual clinical caregiver, and allow linkage to patient data available in routine clinical primary and secondary care datasets.

The Investigator or delegate(s) should immediately countersign and date the form; if a physical ICF is completed, this should then be uploaded to the online trial portal for inclusion in the Investigator Site File (ISF). A copy of the ICF will be given to the participant; where this is an electronic ICF it will automatically be emailed to their nominated email address. Details of the informed consent discussions will be recorded in the participant's medical notes, including date of discussion, the name of the trial, summary of discussion, version number of the PIL given to participant, and version number of ICF signed and date consent received.

### 5 STUDY INTERVENTION

#### 5.1 Pharmacometabolomic treatment approach

To identify markers of hypertension treatment response, machine learning-based data analytics have been applied by a team at the University of Birmingham to multiple metabolomic and clinical datasets from pan-European patient cohorts across the HYPERMARKER consortium. The analysis considers which of the major classes of antihypertensive drugs a person may best respond to, based on their metabolomic and clinical profile. The output from this analysis forms the basis of the information provided to Clinical Investigators for the pharmacometabolomic approach. Clinical Investigators may then choose to consider this additional information when making their own informed clinical prescribing decision. The output will not be provided directly to participants.

The range of drugs considered by the pharmacometabolomic approach are well-established hypertension medications that are the core basis of usual standard of care for hypertension treatment. All treatment decisions are made by the local Investigator who should be clinically-qualified with a licence to practice and prescribe.

To ensure clarity that the prescription a participant receives is the ultimate decision and responsibility of the Clinical Investigator throughout the trial, they will be presented with the following disclaimer statement when presented with the output of the pharmacometabolomic approach:

*"Your patient has had a metabolomic profile related to hypertension medications, and the pharmacometabolomic output provided below combines this with clinical information on your patient to provide a range of potential options. The aim of HYPERMARKER is to support your clinical decision on which blood pressure-lowering medication to use. It is not designed to replace your clinical decision-making. The below output only considers major classes of antihypertensive drugs and may not be appropriate for your specific patient. Please prescribe according to your clinical judgement - we would be grateful for feedback if this deviates from the pharmacometabolomic approach. This is a proof-of-concept pragmatic trial, and the pharmacometabolomic approach is not definitive but will be updated throughout the trial period."*

#### 5.2 Iterative refinement of the model

The pharmacometabolomic approach will be iterated and refined during the trial as additional metabolomic and clinical data from pre-existing cohorts and trial participant data is obtained and clinician feedback is received. Participant data will be kept within an anonymised privacy-preserving Findable, Accessible, Interoperable, and Reusable (FAIR) data repository for this use. Iteration will occur throughout the trial so that the output of the pharmacometabolomic approach can be improved in the second phase of the trial.

#### 5.3 Standard of care

Standard of care for this trial is defined according to the 2024 European Society of Cardiology Guidelines for the management of elevated blood pressure and hypertension (***Figure 2***), with combination therapy recommended for most patients.[1]

Relevant drugs utilised in this trial are:

- Angiotensin-Converting Enzyme (ACE) inhibitors: Inhibit the activity of angiotensin-converting enzyme, thus reducing the production of the vasoconstrictor angiotensin II.
- Angiotensin II Receptor Blockers (ARB): Inhibit the activity of angiotensin II.
- Calcium Channel Blockers (CCB): Prevent calcium from entering the cells of the heart and blood vessel walls, resulting in relaxation and dilatation.
- Diuretics: Increase urine production through changes in urinary sodium, thereby reducing fluid volume. Includes thiazides and thiazide-like diuretics which inhibit sodium-chloride transport in the convoluted tubule of the kidney and mineralocorticoid receptor antagonists which inhibit aldosterone in the distal nephron.
- Beta-blockers: Inhibit beta-receptors in the heart and blood vessels, slowing heart rate, reducing cardiac workload and leading to vasodilation.
- Alpha blockers: Inhibit norepinephrine-induced vasoconstriction.

Some therapies have clinical indications beyond hypertension and so should be continued as part of usual care. All treatment decisions are made by the Clinical Investigator with full knowledge of the participants medical history and previous drug intolerances/allergies.

***Figure 2. 2024 European Society of Cardiology Guidelines for the management of elevated blood pressure and hypertension..[1]***

[redacted]

### 6 STUDY ASSESSMENTS AND PROCEDURES

All planned study assessments and procedures for both the intervention arm as well as the control arm are summarised in the Summary of Assessments (Appendix 1).

#### 6.1 Screening procedure

All screening evaluations must be completed and reviewed to confirm that potential participants meet all eligibility criteria. The Investigator will maintain a screening log to record details of all participants screened and to record reasons for screening failure, as applicable.

#### 6.2 Standard clinical care

The standard clinical care of a person with hypertension may include (but is not limited to): blood tests including to monitor medication safety (such as renal function and electrolytes), screening for hypertensive-mediated organ damage and relevant comorbidities, assessment of cardiovascular risk, physical examination and assessment of medical history. This trial is embedded within usual clinical practice and so it is expected that standard clinical care procedures will be/have been completed by the participants' usual treating physician. Where needed, the Investigator may perform or request standard of care procedures in line with local medical practice.

#### 6.3 Medical history

Medical history relevant to the trial will be collected by the Investigator from medical health records and discussion with the participant, and recorded on the electronic case report form (eCRF). If the Investigator requires additional information to conduct the trial, it is the Investigators responsibility to contact the participant's usual physician(s). The eligibility assessments should be documented in the patient's medical record.

#### 6.4 Blood sampling

Blood sampling to assess metabolomic profiles and medications will be conducted at enrolment and optionally at the end of the trial. 4 mL blood samples using EDTA tubes will be collected by a trained healthcare professional using local venepuncture techniques. Post-collection, pressure will be applied to the puncture site, and participants will be observed briefly for any immediate adverse reactions. From these blood samples, a minimum of 250µL of plasma will be obtained. The ICF explicitly consents for participants samples to be sent to a central laboratory for analysis: Biomedical Metabolomics Facility Leiden (BMFL), Leiden University, Netherlands. Local storage at site may be required temporarily prior to sample transport, with storage process and conditions adhering to site standard operating procedures.

#### 6.5 Study application and training

Participation in the trial will require the Viduet Health application to be downloaded on the participant's personal smartphone and/or tablet and a personal account to be created. The primary purpose of the application in the trial is to record home blood pressure measurements. The path for blood pressure measurement to registration in the Conformité Européene (CE) Medical Device Regulation (MDR)-certified platform is fully closed: measurements are stored digitally in the blood pressure monitor and then, using a secure Bluetooth protocol, sent via the patient's mobile phone signal or WiFi through a secure path to EU-based servers in Frankfurt. The application has a notification system to automatically remind patients of their required measurements. Participants can also be contacted by staff if there are any problems or delays to entry. Messages can be sent via the smartphone application by the research team to the participant, with the planned use outlined in sections 3.4 and 3.5. The system has been used in multiple research trials and is in use in primary and secondary healthcare.

Participants will be trained on study procedures and the Viduet Health application during the enrolment visit. In addition, the training received during the visits will be supported by e-learning material available in the Viduet Health study application and platform. It is the responsibility of the Investigator to ensure that a participant is sufficiently trained on all aspects of the clinical trial.

#### 6.6 Home blood pressure monitoring

Blood pressure will be measured using a semi-automatic, validated and CE-marked monitor (Rossmax Z5 monitor). Home blood pressure measurement will be:

- For at least 3 days per monitoring week.
- At least a reading in the morning and the evening on each day.
- Two blood pressure measurements taken at each timepoint, performed 1-2 minutes apart.
- Taken in a quiet room after 5 minutes of rest, with the patient seated and their arm supported.

The Rossmax Z5 monitor has arrhythmia technology embedded within its functionality to detect the heart rhythm disorder atrial fibrillation. This function is not being used for the purposes of this trial. Where the machine reports atrial fibrillation detection, participants will be advised to consult their usual treating physician.

#### 6.7 Patient-reported outcomes

Questionnaires are prompted by text or email invitation and filled out using any electronic device/system (Phone, tablet, laptop or desktop PC) via the REDCap system web-browser. Participants will complete scheduled electronic patient-reported outcomes (ePRO) assessments according to the Summary of Assessments (Appendix). The questionnaires should be completed independently by the participant. Participants receive programmed reminders to complete the questionnaire. In case of premature discontinuation of treatment, participants will be encouraged to still complete all ePRO questionnaires as planned. The following ePROs will be collected via this method:

- <u>Quality of life:</u> The EuroQOL EQ-5D-5L questionnaire asks about current health status, containing 5 questions on walking mobility, self-care, usual activities, pain/discomfort and anxiety/depression, and a visual analogue scale on health status perception. Collection point: Baseline, and following each phase of the trial (approximately week 6 and week 12). Timeframe: Day of completion.
- <u>Healthcare resource utilisation:</u> This questionnaire collects data relevant to direct and indirect healthcare utilisation, including number of hospital visits (inpatient, outpatient and emergency department); the number of primary and community care (general practitioner, nursing and allied health professionals); the number of ambulatory services; medications; diagnostic tests and potential productivity loss due to time off work. Collection point: Baseline and following each phase of the trial (approximately week 6 and week 12). Timeframe: Last 6 weeks or since last ePRO collection (whichever is shorter).
- <u>Adherence questionnaire:</u> Participants will be asked about whether and how many times they have not taken their anti-hypertensive medication, which therapy has been omitted and for what reason, and any perceived adverse reactions from a common list of side-effects as listed on relevant Summary of Product Characteristics for different anti-hypertensive medications. Collection point: Following each phase of the trial (approximately week 6 and week 12). Timeframe: last 2 weeks.
- <u>Socioeconomic status (enrolment only; optional):</u> This questionnaire contains questions on marital status, education level, income, employment status, profession, and number of persons in the household.
- <u>Demographics (enrolment only; optional):</u> This questionnaire collects data on participant characteristics, including country of birth and ethnicity and is combined with the above questionnaire. Ethnic groups will be defined by the 2021 census of England and Wales and are included due to variation in the prevalence of hypertension and its complications across these groups.[25,26]

A dietary intake questionnaire will be completed by participants at site using an electronic device/system (Phone, tablet, laptop or desktop PC). Questionnaires will be hosted by myfood24 (Dietary Assessment Ltd), for sites in the UK, Germany & Spain and Compl-eat (Wageningen University), for sites in the Netherlands and collect data regarding participants food and drink consumption from the previous 24hrs.

Collection point: At metabolomic testing (enrolment visit and optional end of trial visit). Timeframe: 24-hour period prior to blood test.

##### 6.8 Pregnancy testing

Certain anti-hypertensive medications should be avoided in pregnancy, as per normal clinical standard of care. Women of childbearing potential (WOCBP), will be advised of the risk from certain anti-hypertensive medications within pregnancy and advised of the importance of avoiding pregnancy during the trial. Where WOCBP cannot verbally exclude whether they could be pregnant at enrolment, a urinary pregnancy test will be offered. In the event a participant becomes pregnant during the trial they will be withdrawn.

As per the Heads of Medicines Agencies Clinical Trials Facilitation and Coordination Group (2020), the definition of WOCBP that Investigators should use is as follows: "Fertile, following menarche and until becoming post-menopausal, unless permanently sterile. Permanent sterilisation methods include hysterectomy, bilateral salpingectomy and bilateral oophorectomy. A postmenopausal state is defined as no menses for 12 months without an alternative medical cause. A high follicle stimulating hormone level in the postmenopausal range may be used to confirm a postmenopausal state in women not using hormonal contraception or hormonal replacement therapy. However, in the absence of 12 months of amenorrhea, confirmation with more than one follicle stimulating hormone measurement is required."

##### 6.9 End of trial definition and subsequent management

The end of study for each participant is defined as the date the participant completes their last patient-reported SBP or ePRO. Following the end of trial participation, continuation of any blood pressure treatment will be determined by each participants' usual treating physician, defaulting to usual standard of care. The 'overall end of trial' is defined as the completion of the last patient-reported SBP or ePRO of the last participant.

##### 6.10 Discontinuation/withdrawal

Participants can leave the study at any time and for any reason without any consequences; their Investigator and primary caregiver will be notified and provided with a summary of any blood pressure recording data up to the date of withdrawal.

An Investigator may deem it necessary to withdraw a participant from the trial if any clinical adverse event, laboratory abnormality, or other medical condition or situation occurs such that continued participation in the trial would not be in the best interest of the participant.

All study withdrawals, including the date and reason, should be recorded by the Investigator in the eCRF and in the participant's medical records.

#### 7 OUTCOMES AND ANALYSIS

##### 7.1 Primary outcome

Change in home SBP will be derived from all available patient-measured SBP recordings in the study smartphone application, comparing the intervention and standard of care groups at the end of the first phase of the trial. This includes 1-week of monitoring after enrolment (anticipated minimum of 12 recordings) and at least 4-weeks of monitoring after therapy change (anticipated minimum of 48 recordings).

##### 7.2 Secondary outcomes

The following secondary outcomes will compare the pharmacometabolomic-guided drug class approach and standard of care groups at the end of the first phase of the trial:

1. Proportion of participants achieving a target home SBP of 120–129mmHg using the average of the final 3 days of blood pressure measurements.
2. Proportion of participants reporting any treatment-related adverse effects compiled from the Summary of Product Characteristics from the different classes of anti-hypertensive medications.
3. Proportion of participants reporting withdrawal of an anti-hypertensive medication.
4. Proportion of participants reporting ≥90% adherence to prescribed anti-hypertensive medication.
5. Rate of change in home SBP using all available SBP measurements, averaged per week.
6. Change in home diastolic blood pressure derived from all available blood pressure recordings.

The following secondary outcomes will separately compare the original intervention, updated intervention and standard of care groups at the end of the second phase of the trial:

7. Change in home SBP using all available SBP measurements, comparing the iterated pharmacometabolomic approach versus the initial pharmacometabolomic approach, and the iterated pharmacometabolomic approach versus initial standard of care.
8. Patient-reported treatment-related side effects, comparing the iterated pharmacometabolomic approach versus the initial pharmacometabolomic approach, and the iterated pharmacometabolomic approach versus initial standard of care.

The following outcomes will apply across the phases of the trial comparing any pharmacometabolomic approach versus standard of care:

9. Proportion and number of serious adverse events, including all-cause hospitalisation and death.
10. Proportion and number of healthcare utilisation events, including details on hospitalisation (frequency, cause, type [outpatient, emergency, admission] and length of stay) and primary care interaction (frequency, cause and type [doctor, nurse, other allied health professional]).
11. Patient-reported quality of life using the EQ-5D-5L summary index score and visual analogue scale.

The following outcomes are unrelated to the randomised group:

12. Change in metabolomic profile from baseline to (optional) follow-up blood sample, stratified by class of anti-hypertensive medication (exploratory).
13. Association between dietary intake, metabolomic profile and blood pressure response to prescribed antihypertensive treatment (exploratory).

#### 7.3 Health economic outcomes

The economic evaluation will be divided into two separate but complementary analyses:

Health resource utilisation (trial-based analysis)**:** This analysis focuses on health resource utilisation based on the trial results. Specifically, a comparative analysis of resource utilisation between the two arms will be conducted. Baseline characteristics of patients in the intervention and standard-of-care groups according to intention-to-treat will be summarised using data entered by patients via REDCap ePROs. Healthcare resource use will be compared between the two groups using two-sample t-tests and Chi-squared tests for continuous and categorical variables, respectively. The key economic evaluation outputs will include the net monetary benefit, incremental cost-effectiveness ratios, the cost-effectiveness planes and the cost-effectiveness acceptability curves, discounted at recommended values. To address uncertainty, the analysis will include both univariate deterministic sensitivity analysis and probabilistic sensitivity analysis.

Health resource utilisation (care pathway mapping):

This analysis examines healthcare resource utilisation data collected during the care pathway mapping. Care activities will be systematically identified and sequenced, with resource use measured and valued by the London School of Economics (LSE) health economics research team.

Participants from selected study sites who have signed the relevant optional section of the informed consent form may be invited to partake in a semi-structured and open-ended interview with the research team. A total of 8-12 participants will be required per selected site. Interviews will be conducted at the trial site within the trial period. Researchers will use an iterative approach to develop an initial prototype of the care pathway. This process will identify critical decision points, initial diagnosis, care gaps, and patient flow through the healthcare system.

The interviews will follow a structured topic guide covering three key areas: sociodemographic characteristics, patient comorbidities, and key decision points in diagnosing and managing hypertension. Participants can decide whether to extend or shorten the duration of their sessions based on their preferences.

The research team will use semi-structured and open-ended questions to explore patient experiences in depth. Interviews will be audio recorded, and the research team will transcribe interviews verbatim and analyse them using thematic analysis, applying open and axial coding methods from grounded theory. This process will generate a visual process map that represents the current care pathway and reveals how different activities in hypertension diagnosis and management are connected. The team will analyse data iteratively, using insights from early interviews to refine subsequent ones. Thematic coding will identify key events and linkages in the patient care journey, helping to highlight gaps and inefficiencies in the healthcare system. Participants who complete interviews may be invited for a follow-up appointment with the LSE research team to further explore interview topics. These may be conducted in person, online or via telephone.

#### 7.4 Sample size calculations

Multiple repeated measures can achieve much greater power than a single time-point measurement;[27] using each participant as their own control reduces subject-to-subject variability explained by anything other than the effect of the treatment under study. The sample size of 400 patients using home SBP provides 99% power to detect a 1.5mmHg difference in SBP between the pharmacometabolomic approach and standard of care groups. Sample size calculations are based on the expected minimum of 12 baseline SBP measurements and 48 follow-up measurements by the end of the first phase, and assume a 2-sided alpha of 0.05, baseline SBP of 145 mmHg (SD 20), with correlation between SBP measurements of 0.7.[28] Power is retained at >85% for a 2.5mmHg difference in home SBP even if only 3 baseline and 12 follow-up measurements are recorded (with correlation 0.5 and 2-sided alpha 0.05).

#### 7.5 Statistical analysis

A comprehensive Statistical Analysis Plan (SAP) detailing all analyses will be developed and signed off prior to final data lock and any analysis of trial outcomes. Any deviations from the SAP after this point will be documented, explained in the final trial report and considered exploratory. The primary analysis will follow the intention-to-treat (ITT) principle, analysing all participants in their originally assigned groups regardless of compliance with the allocated group, with no imputation for any missing data. Sensitivity analyses will include ‘as-treated’ and ‘per-protocol’ analyses. A p-value <0.05 will be considered statistically significant.

The primary outcome of change in home SBP using multiple repeated measurements will be analysed using generalised linear models generated using a random-effects estimator and exchangeable correlation matrix. These account for all available SBP readings, accommodate variability in measurement frequency and intervals between participants, and account for repeated measures correlation. Models will include adjustment for variables used in the randomised stratification (age and site), as well as gender, prior blood pressure treatment and relevant comorbidities.

Secondary outcomes will be analysed using the same methods as described above (for continuous repeated outcomes), adjusted mean difference (for other continuous outcomes), logistic or log-binomial regression (for categorical outcomes), chi-squared or Poisson regression (for count data), and incidence rate ratio (for adverse events).

### 8 STORAGE AND HANDLING OF HUMAN TISSUE

#### 8.1 Collection and handling of human tissue

Blood samples will be obtained by a clinically-trained member of the local research team (as outlined in section 6.4) and submitted to the site-specific laboratory as per local guidelines and SOPs, ensuring that all specimens are accurately labelled and handled appropriately. Samples for metabolomic profiling will be prepared and may be temporarily stored at their respective sites before being transported for analysis to the Biomedical Metabolomics Facility Leiden (BMFL), Leiden University, the Netherlands, as outlined in the agreed lab manual. Upon arrival of samples at BMFL, individual items will be inspected, and the Samples Receiving Form (F1-SP 20.01) filled in. The Sample Shipment Form (F3-SP 20.01) will be checked against the delivered samples for consistency. When samples will not be analysed immediately, they will be stored at -80°C (or as otherwise required). Each sample will have a unique sample number.

#### 8.2 Storage and disposal of human tissue

Measured samples will be stored at 4C or -80C until 30 days after reporting of the metabolomics results after which the original samples will be disposed of in accordance with relevant regulations. All samples are accessible by BMFL personnel prior to disposal.

### 9 SAFETY AND OVERSIGHT

HYPERMARKER is a proof-of-concept strategy trial that may affect the order that existing and approved classes of medications for hypertension are used in individual patients. All medications used by Clinical Investigators should be recommended in international hypertension guidelines (e.g., European Society of Cardiology[1]) as well as local formularies at each clinical site. Clinical Investigators should only align their prescribing practice with the output of the pharmacometabolomic approach if they deem it to be clinically appropriate and in line with licensed indications for their patient.

The study is not a Clinical Trial of an Investigational Medicinal Product (non-CTIMP designation) and is therefore not subject to EU or UK regulations for medicinal products. Typical adverse events from antihypertensive therapy will be recorded from participants to aid in future iteration of smart treatment selection approaches. As per Good Clinical Practice (GCP) requirements, serious adverse events (SAEs) will still be recorded during the research period, even though in this short duration non-CTIMP study they are likely to be unrelated to the research question.

#### 9.1 Expected adverse events

Adverse events will be collected directly from the ePRO questionnaires. Most adverse events/adverse reactions that occur in this trial will be expected treatment-related undesirable effects from drugs used for hypertension in routine clinical practice. Common side effects (collated from the Summary of Product Characteristics) for frequently-used anti-hypertensive drugs are: abdominal pain, angioedema, blood glucose abnormalities, chest pain, confusion, cough, dizziness & syncope, dyspnoea, electrolyte disturbance, fatigue, flushing, gastrointestinal disturbance, gynaecomastia, headache, hepato-biliary disorders, hypersensitivity, hypotension, incontinence, infections (respiratory tract & urine), joint pain, lipid disorders, mood disorders, muscle pain and spasm, oedema/ankle swelling, palpitations & arrhythmia, paraesthesia, rash, Raynaud's, renal impairment, sexual dysfunction, sleep disturbance, vasculitis and visual disturbance.

Fluctuations in blood pressure readings are an expected part of natural variability in physiology and recording measures. Blood pressure readings deemed clinically significant will automatically be flagged for review by the study team, with participants asked to repeat their measurement and seek clinical attention as needed.

##### 9.2 Serious adverse events

An SAE is defined as an untoward occurrence that: a) results in death; b) is life-threatening; c) requires hospitalisation or prolongation of existing hospitalisation; d) results in persistent or significant disability or incapacity; e) consists of a congenital abnormality or birth defect; or f) is otherwise considered medically significant by the Investigator. SAEs that occur during the study period will be reported by Clinical Investigators using a specific SAE eCRF form. A paper-based option is also available in case electronic systems are temporarily not functioning. All SAEs will be followed up until resolution, or participant completion in the study, whichever is earlier.

In line with non-CTIMP studies embedded in routine clinical practice, only SAEs related to administration of any of the research procedures and deemed as an unexpected occurrence by the Chief Investigator or delegate will be reported in line with the sponsor's safety reporting procedures to the REC and Sponsor (within 15 days of the Chief Investigator or delegate becoming aware of the event).

##### 9.3 Trial Management Group

The Trial Management Group (TMG) will comprise the core team and Principal Investigators as listed on the key study contacts page, as well as administrative staff and ad-hoc staff needed to deliver the trial. The TMG will convene at regular intervals and be responsible for the day-to-day running and management of the trial.

##### 9.4 Joint Trial Oversight Committee

A joint oversight committee comprising a Trial Steering Committee (TSC) and Data Monitoring Committee (DMC) will be engaged for this trial. The role of the TSC is to provide the overall supervision of the trial. The TSC will monitor trial progress and advise on scientific credibility. The TSC will consider and act, as appropriate, upon the recommendations of the DMC whose role is to oversee the safety of participants in the trial. The TSC and DMC will operate in accordance with their trial-specific charters, based on the DAMOCLES recommendations.[29] Due to the short nature of the trial, virtual meetings of the joint oversight committee are expected to occur once prior to the trial opening to enrolment, and once after participant recruitment has completed, with additional meetings ad-hoc as needed.

#### 10 ETHICAL AND REGULATORY CONSIDERATIONS

##### 10.1 Assessment and management of risk

To reduce the burden on participants, the trial will be embedded in routine clinical practice with enrolment planned to take place during secondary care encounters. Trial procedures after enrolment can take place remotely to further reduce this burden. The information provided to prospective participants has been co-developed with a PPI group with representatives from all four trial site countries to ensure potential participants are provided adequate information on which to base their informed decision when providing consent.

This trial is testing a prescribing strategy for antihypertensive drug class, with standard of care versus a smart pharmacometabolomic-guided approach. The latter uses machine learning-based data analytics to generate output, which is outlined in the participant information leaflet. This analysis will be conducted by the sponsor institution (University of Birmingham) to limit data transfers and ensure the safe processing of participant data. All blood pressure medications to be used are licenced and part of standard of care, and the trial does not require any standard of care procedures to be withheld throughout the trial. The output of the smart pharmacometabolomic approach is not directly involved in the prescribing process for a patient. The Clinical Investigator will be presented with the output, but retains the decision for treatment prescription regardless of randomised allocation. This study is not a medical device trial, and to ensure clarity that the prescription a participant receives is the decision and responsibility of the clinical investigator throughout the trial, they will be presented a disclaimer statement as previously described in section 5.1. Clinical Investigators will have access to the clinical data through the REDCap system and Viduet platform on which the output of the pharmacometabolomic approach is based when making their own informed prescribing decision. Within REDCap, Investigators will record medications prescribed and provide feedback to aid in future development and, if this proof-of-concept trial is successful, inform subsequent medical device development trials.

Participants will undergo an enhanced level of self-monitoring with this burden mitigated by the provision of a blood pressure monitor and mobile app to allow at-home monitoring. An enhanced level of monitoring also mitigates the risk of blood pressure fluctuations during medication changes with in-built safety alerts within the app alerting the clinical team of significant recordings. The patient's usual doctor (e.g. General practitioner) will also be informed of their involvement and, following the end of the participants time in the trial will receive a copy of the patient's average trial blood pressure and treatments to ensure safe handover of care. Conversely, participants stand to gain potential benefits from a personalised treatment approach, including improved blood pressure control, less side effects and better overall health outcomes. The group-relatedness aspect is addressed through randomisation stratification and the two-phase design, ensuring a balanced distribution of key variables and providing all participants with the opportunity to benefit from the personalised approach over the course of the study.

Participants will face a burden associated with providing blood samples and completing electronic questionnaires. Only one mandatory and one optional blood sample are required to mitigate this burden, and the electronic questionnaires can be completed at home with adequate training and support provided by research staff at each site for at-home device use and data collection.

#### 10.2 Ethical approval

The trial will be performed in accordance with the principles of the Declaration of Helsinki and conducted in accordance with all applicable research governance frameworks statutory instruments (which includes the Medicines for Human Use Clinical Trials 2004 and subsequent amendments, the latest Data Protection legislation, the Human Tissue Act 2008, EU Clinical Trials Directives and Clinical Trials Regulation and amendments, and Guidelines for GCP.

Following Sponsor approval and before the start of the project, a favourable opinion will be sought from a research ethics committee for the protocol, informed consent forms and other relevant documents.

#### 10.3 Regulatory review and compliance

Before any site can enrol participants into the project, the CI/Principal Investigator or designee will ensure that appropriate approvals from participating organisations are in place. Specific arrangements on how to gain approval from participating organisations are in place and comply with the relevant guidance. For any amendment to the project, the CI or designee, in agreement with the sponsor will submit information to the appropriate body in order for them to issue approval for the amendment. The CI or designee will work with sites (R&D departments at NHS sites as well as the project delivery team) so they can put the necessary arrangements in place to implement the amendment to confirm their support for the project as amended. The University of Birmingham's Clinical Research Compliance Team may carry out compliance visits to monitor adherence with applicable standards and regulations. Due to the short length of this trial, the CI or delegate will only send an 'End-of-Trial' notification and final report to the Sponsor and approval bodies.

#### 10.4 Site set-up and monitoring

All members of the site research team will be required to sign a site signature and delegation log and have completed the site initiation and GCP training. The PI is responsible for creating and updating an

Investigator Site File containing essential documentation, instructions, and other documentation required for the conduct of the trial. The CI or delegate should be informed immediately of any change in the site research team. As this trial is embedded in routine clinical care, no specific monitoring is planned. Monitoring may be triggered, for example by poor eCRF return, poor data quality, or excessive participant withdrawals or deviations. Investigators are required to allow the core trial team access to source documents as needed to address these issues. In case of audit or inspection by the Sponsor or regulatory bodies, the PI will permit trial-related monitoring and quality checks at their site, providing direct access to source data and documents, in line with consent.

#### 10.5 End of study and archiving

The 'overall end of trial' is defined as the completion of the last patient-reported SBP or ePRO of the last participant.

Archiving will be authorised by the Sponsor following submission of the end-of-trial report, and will include relevant trial documents, the trial database and all essential material for a minimum of 10 years, or according to the sites local standard operating procedure after completion of the trial. Archiving and destruction of documents will follow the Sponsor's Standard Operating Procedures.

#### 10.6 Protocol Deviations

A study related deviation is a departure from the ethically-approved study protocol, or other study document or process (e.g., consent process or administration of study intervention), or from GCP or any applicable regulatory requirements. Accidental protocol deviations can happen at any time. They must be adequately documented on the relevant forms, filed in the trial master file and reported to the CI and Sponsor immediately. Significant deviations from the protocol which are found to frequently recur are not acceptable; these will require immediate action and could potentially be classified as a serious breach. Serious breaches (which are deemed as likely to affect to a significant degree the safety or physical/mental integrity of the participant, or scientific value of the research) should be notified to the Sponsor within 1 working day. In collaboration with the CI, the serious breach will be reviewed by the Sponsor and, if appropriate, the sponsor will notify the REC and other regulatory bodies without undue delay, and no later than seven days after becoming aware of the breach.

#### 10.7 Indemnity/insurance

The University of Birmingham has in place indemnity coverage for this trial which provides cover to the University for harm which comes about through negligence by the University or its staff in relation to the design or management of the trial, and may alternatively, and at the University's discretion, provide cover for non-negligent harm to participants. The University of Birmingham is independent of any pharmaceutical company, and as such it is not covered by the Association of the British Pharmaceutical Industry guidelines for participant compensation. With respect to the conduct of the trial at the Clinical Sites and other clinical care of the patient, responsibility for the care of the patients remains with the organisation responsible for the Clinical Site and is therefore indemnified through their local policies.

#### 10.8 Peer review and public involvement

Expert, independent review of the trial programme has been undertaken as part of the funding application to the EU Horizon and UKRI schemes (HYPERMARKER 101095480). This protocol has been reviewed within the Sponsor's institution (University of Birmingham) and sites (University Medical Center Utrecht, INCLIVA Instituto de Investigación Sanitaria, University Medical Center Hamburg-Eppendorf and University Hospitals Birmingham NHS Foundation Trust).

This protocol and patient-facing material have been developed in conjunction with a Patient and Public Involvement (PPI) team, in line with the PPI-POSITIVE approach[30] and CODE-EHR framework.[31]

### 11 DATA AND DATA PROTECTION

All Investigators and trial site staff must comply with the requirements of the General Data Protection Regulation (GDPR) with regards to the collection, storage, processing and disclosure of personal information and will uphold its core principles. Access will be limited to the minimum number of individuals necessary for quality control, audit and analysis. The controller of the data is the University of Birmingham, and all staff are expected to comply with this institution's Standard Operating Procedures.

In brief, all data will be used in line with the UK Data Protection Act 2018, consistent with EU GDPR. For example, the principles of: (1) Fair, lawful and transparent use by only using anonymised data for analysis; (2) Explicit use of this data for the purposes of health improvement in specified patient subgroups; (3) Relevant and limited use of data to what is necessary to answer the research questions; (4) Application of established data pipelines to ensure accuracy, and identify and rectify anomalies; (5) Keeping data for no longer than is necessary and permit collaboration/data sharing with other research groups, where applicable, to ensure the full extent of value from the data obtained; (6) Handling data in a way that ensures security and prevents loss or misuse; and (7) Technical and organisational procedures in place to ensure accountability, in addition to PPI input on research questions and data use.

#### 11.1 Source documents

Source documents for this study will include hospital records, procedure & investigation reports and specific data collection forms. These documents will be used to enter data directly into the online eCRF. Data reported on the eCRF that are derived from source documents must be consistent with the source documents, or any discrepancies must be explained. All documents will be stored safely in confidential conditions. On all study-specific documents other than the signed consent, the subject will only be referred to by their individual participant identification code.

#### 11.2 Case report forms

HYPERMARKER will utilise the Research Electronic Data Capture (REDCap) system, originally developed by Vanderbilt University with ongoing support from the US National Institutes of Health. REDCap is a browser-based data capture software, supported by an experienced local team at the University of Birmingham. The system as deployed at the University of Birmingham is compliant with the Data Protection Act and GDPR, and includes access restricted to nominated study staff, with clear audit trails for data/user monitoring. User rights per project are governed by the CI or nominated REDCap administrator. All access, changes and addition details are logged within a project's logging section.

REDCap is operated across two virtual servers hosted on the University of Birmingham network: a web application server and a MySQL database server. All University virtual severs are built to a secure standard. Daily backups of the server infrastructure are taken to allow fall backs to previous versions if required. A documented build process for installing REDCap and all security settings is followed by local staff. The University servers sit behind a site firewall that helps protect access. The web application server has a secure connection through a specific firewall rule to the MySQL database server. The authentication to REDCap is via a university user account, along with utilising REDCap's User allow list. An administrator must provide access to REDCap and a secure verification process is required before users can log on. Regular server security scans and reports are produced to identify any missing security patches. The report is emailed to system administrators with the relevant information for patches, upgrades or bug fixes. An external website security scan is checked via a third-party website, the results are stored and can be viewed upon request. The REDCap database is backed up via the College Backup system using CommVault software. Backups occur daily, weekly and monthly and are stored in an offsite fireproof safe.

All incoming data in REDCap gets intentionally filtered, sanitised, and escaped. This includes all data submitted in an HTTP Post request and all query string data found in every URL while accessing REDCap, among other modes through which user-defined data gets submitted in the application. Server environment variables that are vulnerable to forgery by users are also checked and sanitised. All user submitted data is properly filtered for any possibly harmful mark-up tags (e.g. <script>) and then escaped before ever being displayed on a web page within the application. SQL queries sent to the database server from REDCap are all properly escaped before being sent. If any values used in an SQL query originated from user-defined values, they would have already been sanitised beforehand as well, as described above. User-defined data used within SQL queries also have their data type checked to prevent any mismatching of data types (e.g. making sure a number is really a number). These processes of sanitisation, filtering, data type checking, and escaping all help to protect against methods of attack, such as Cross-Site Scripting and SQL Injection. To specifically protect against Cross-Site Request Forgery, REDCap utilises a "nonce" (a secret, user-specific token) on every web form used in the application. The nonce is generated anew on each web page as the user navigates within REDCap during a session.

#### 11.3 Viduet health application

The primary purpose of the Viduet smartphone application within the trial is to collect and record participants data generated from the Rossmax Z5 blood pressure monitor. It may also be used to prompt participants to take their measurements, for the research team to send messages to participants and for safety alerts (e.g. significantly high blood pressure recordings). Participants will need to input their personal details to create a secure Viduet personal account at the start of the trial.

Viduet has an ISO 27001 certified Information Security Management System which includes protocols/procedures/controls about information security and confidentiality that employees of Viduet abide by. Participant data is stored on encrypted databases. The participant medical data acquired (e.g., blood pressure data) cannot be altered by anyone and is stored in Viduet 'as is'. Viduet employee access to participant data is restricted to only those who need access for certain activities (such as participant support).

Each trial site will have access to data for participants recruited at their respective site via the online Viduet Health clinical portal. Pseudonymised home blood pressure data will be exported and securely transmitted multiple times during and at the end of study to the University of Birmingham for the iterative refinement of the pharmacometabolomic approach, as outlined in section 5.2 and for analysis of the trial results.

After the study, data held within Viduet Health will be controlled by participants in line with Viduet Health's data policy. In view of this, participants will be sent a message following the end of study for each participant to inform them of how they can choose for their data to be destroyed from the Viduet database. Data exported to the University of Birmingham will be held within the trial databases and handled at the end of trial as outlined in section 10.5.

#### 11.4 Collection of dietary information

Data will be collected from a validated dietary questionnaire hosted by either the myfood24 (Dietary Assessment Ltd) or Compl-eat (Wageningen University) platforms, to account for the availability of the languages and dietary information required for respective sites. These will be completed by participants at site using an electronic device (smartphone, tablet or computer). Only the participants study ID will be required, no personal identifiable data will be entered. Dietary data will then be transferred to the University of Birmingham.

#### 11.5 Metabolomic analysis of blood samples

Metabolomics data from blood samples will be immediately saved on the PC that is connected to the liquid chromatography–mass spectrometry instrument and backed up to the safe data storage at the (BMFL) of Leiden University, Netherlands. Data back-ups are synchronised daily to a co-location in Delft, Netherlands, with maintenance and responsibility by the IT department of Leiden University. In case of insufficient capacity, additional storage can be purchased from SURFsara, a national Netherlands high-performance computing solution. Data will be made available, freely or upon request, as soon as possible with respect to legal, commercial, ethical or other pressing considerations as determined by the Data Protection Officers (DPOs) of the participating organisations. The DPOs are responsible for monitoring compliance with relevant national and international data privacy and data protection regulation, such as EU GDPR.

##### 11.6 Health economic data

Pseudonymised data provided to the health economics team will reside within secure servers at the London School of Economics (LSE) Primary Data Centre. LSE encrypts the data link between its on-campus firewalls and its firewalls hosted within its own equipment in the Data Centre; there is no facility for the traffic to be intercepted and decrypted. LSE encrypts the traffic using AES 256 between client machines and servers storing the data. All ingress to and egress from the LSE network is controlled by use of next-generation firewalls which are Common Criteria EAL4 compliant. Access to LSE resources is governed by the stringent Access Control Policy of the LSE.

Data gathered from the Health resource utilisation (care pathway mapping) process will be stored, processed and managed within the boundaries of LSE's Information Security Policy. Research data will be kept and registered for at least five years with LSE Library repository and managed according to LSE institutional guidelines on confidentiality, anonymisation, and access restrictions.

##### 11.7 Trial master file

The clinical trial master file shall at all times contain the essential documents relating to the clinical trial, which allow verification of the conduct of the trial and quality of the data generated. The content of the clinical trial master file shall be archived in a way that ensures it is readily available and accessible, upon request, with any medical files of participants archived in accordance with national law.

##### 11.8 Research frameworks

The HYPERMARKER project adheres to quality and transparency frameworks for development of artificial intelligence algorithms[32] and the CODE-EHR standards for use of structured healthcare data in research[31]. This protocol was developed in accordance with the Standard Protocol Items for Randomized Trials (SPIRIT) statement,[33] the SPIRIT-PRO extension[34] and the SPIRIT-Outcomes extension[35]. Trial results will be reported in accordance with the CODE-EHR standards[31] and CONSORT-AI reporting guidelines.[36]

#### 12 DISSEMINATION POLICY

No individual participant data will be disseminated. Publications and presentations relating to the main trial, including abstracts, will summarise group data. All findings of clinical relevance will be submitted to a suitable medical journal for publication after peer review. The PPI team will provide a short lay summary of results that will be published alongside the scientific paper. Named authors on any publication must satisfy the International Committee of Medical Journal Editors (ICMJE) criteria for authorship (contribute to drafting of the article or revision for important intellectual content), provide timely approval of the final version to be published, and supply detailed statements on any potential conflict of interest or financial relationships. Members of the research team who do not fulfil ICMJE criteria for authorship will be listed as collaborators.

### 14 APPENDICES

#### 14.1 Appendix 1 - Schedule of activities

| Procedure | | Enrolment | Between phase 1 and 2 | End of Trial |
|---|---|---|---|---|
| Assessment of eligibility criteria | | ● | | |
| Informed consent obtained | | ● | | |
| Blood sample for metabolomics | | ● | | ● (Optional) |
| Review of medical & medication history | | ● | ○ | |
| Physical exam | Office blood pressure & Heart Rate | ● | | ● (Optional) |
| Physical exam | Body weight & height | ● | | |
| Study device & app installation & training | | ● | | |
| Randomisation | | ● | | |
| Electronic patient-reported questionnaires | Socioeconomic status & demographics | ○ (Optional) | | |
| Electronic patient-reported questionnaires | 24-hr dietary intake | ● | | ● (Optional) |
| Electronic patient-reported questionnaires | Quality of life (EQ-5D-5L) | ○ | ○ | ○ |
| Electronic patient-reported questionnaires | Adherence & adverse reactions | | ○ | ○ |
| Electronic patient-reported questionnaires | Healthcare utilisation | ○ | ○ | ○ |
| At-home blood pressure measurements | | ○ | ○ | ○ |
| Review of participant blood pressure data by investigator # | | ○ | ○ | |
| Review and prescription of blood pressure medication by Investigator # | | ○ | ○ | |

*ePRO, Electronic Patient-reported Outcomes*
*Procedures indicated by ● indicate in-person collection. Procedures indicated by ○ will be conducted remotely.*
*# Review outside of these timepoints may occur as directed by the built-in portal alerts (or participant enquiries) or to maintain participants safety.*

#### 14.2 Appendix 2 – amendment history

| **Protocol Amendments** | | | | |
|---|---|---|---|---|
| The following amendments and/or administrative changes have been made to this protocol since the implementation of the first approved version: | | | | |
| **Amendment number** | **Date of amendment** | **Protocol version number** | **Type of amendment** | **Summary of amendment** |
